# Causal Estimands for Analyses of Averted and Avertible Outcomes due to Infectious Disease Interventions

**DOI:** 10.1101/2024.07.24.24310946

**Authors:** Katherine M Jia, Christopher B Boyer, Jacco Wallinga, Marc Lipsitch

## Abstract

During the coronavirus disease (COVID-19) pandemic, researchers attempted to estimate the number of averted and avertible outcomes due to non-pharmaceutical interventions or vaccination campaigns to quantify public health impact. However, the estimands used in these analyses have not been previously formalized. It is also unclear how these analyses relate to the broader framework of direct, indirect, total, and overall causal effects of an intervention under interference. In this study, using potential outcome notation, we adjust the direct and overall effects to accommodate analyses of averted and avertible outcomes. We use this framework to interrogate the commonly-held assumption in empirical studies that vaccine-averted outcomes via direct impact among vaccinated individuals (or vaccine-avertible outcomes via direct impact among unvaccinated individuals) is a lower bound on vaccine-averted (or -avertible) outcomes overall. To do so, we describe a susceptible-infected-recovered-death model stratified by vaccination status. When vaccine efficacies wane, the lower bound fails for vaccine-avertible outcomes. When transmission or fatality parameters increase over time, the lower bound fails for both vaccine-averted and -avertible outcomes. Only in the simplest scenario where vaccine efficacies, transmission, and fatality parameters are constant over time, outcomes averted via direct impact among vaccinated individuals (or outcomes avertible via direct impact among unvaccinated individuals) is shown to be a lower bound on overall impact on vaccine-averted (or -avertible) outcomes. In conclusion, the lower bound can fail under common violations to assumptions on constant vaccine efficacy, pathogen properties, or behavioral parameters over time. In real data analyses, estimating what seems like a lower bound on overall impact through estimating direct impact may be inadvisable without examining the directions of indirect effects. By classifying estimands for averted and avertible outcomes and examining their relations, this study improves conduct and interpretation of research evaluating impact of infectious disease interventions.

## 1. INTRODUCTION

Recently, researchers have estimated the number of COVID-19 deaths (or infections) averted by vaccination campaigns in the United States,^1,2^ Israel,^3,4^ Chile,^5^ Brazil,^6^ and Japan.^7^ Similarly, other studies, including our own,^8^ have attempted to estimate vaccine-avertible deaths—defined as the number of deaths that could have been averted by vaccination, but were not because of a failure to vaccinate the unvaccinated.^8,9^ Most empirical studies quantify direct protection conferred by vaccination among vaccinated individuals, but they typically assume the overall impact of vaccination is larger due to indirect protection.^4–6^ Indeed, these studies often claim that their estimates represent a lower bound on overall impact of vaccination, although this claim has not been carefully verified.

To support this claim, many of these studies draw, either implicitly or explicitly, on the causal effect framework developed by Halloran and Struchiner,^10^ in which the overall effect of vaccines can be decomposed into direct and indirect components.^11^ However, the Halloran and Struchiner framework has not yet been formally extended to cover the specific estimands targeted in vaccine-averted and avertable analyses, which estimate *impact*. Existing effect estimands are defined by contrasts of individual risk^12,13^ and have been used to estimate vaccine efficacy in clinical trials.^14,15^ However, observational studies, post-licensure studies, and policy-makers are often equally or more interested in quantifying public health *impact* of vaccination in terms of counts^16^ such as the number of infections, hospitalizations, and deaths in a group of individuals, instead of risk in each individual.

Motivated by recent empirical studies on vaccine-averted and -avertible COVID-19 deaths, this article seeks to fill these gaps by: (1) Clearly defining *impact* estimands as corollaries of direct and overall effect estimands for averted and avertible outcomes (i.e., counts), and (2) determining the conditions under which direct impact is a lower bound on overall impact. To ground our discussion, we introduce a susceptible-infected-recovered-death (SIRD) model stratified by vaccination status to investigate direct and overall impact of vaccination under different scenarios. Section 2 summarizes the definitions of direct, indirect, total, and overall effects introduced by Hudgens and Halloran,^12^ and provides an alternative partitioning of overall effect into components that will align better with the estimands targeted by vaccine-averted and avertible analyses. Section 3 proposes count outcome corollaries for direct and overall effects, shows how they map onto estimands for vaccine-averted and -avertible analyses, and formalizes the claim that direct impact constitutes a lower bound on overall impact. Section 4 outlines a transmission model to simulate vaccine-averted and -avertible outcomes. Section 5 examines the conditions under which outcomes averted via direct impact among vaccinated individuals (or outcomes avertible via direct impact among unvaccinated individuals) is or is not a lower bound on overall impact on vaccine-averted (or -avertible) outcomes.

## 2. DIRECT, INDIRECT, OVERALL, AND TOTAL EFFECTS

Hudgens and Halloran^12^ previously defined causal estimands for direct, indirect, overall, and total effects in the two-stage randomized trial, as summarized below.

### Setup and notation

Consider a two-stage randomized trial with *m* groups indexed by *i* = 1, … , *m*, such that each group consists of *N* individuals indexed by *j* = 1, … , *N* with a large group size *N*. All groups are assumed to be of same size *N*. Partial interference is assumed: Individuals make contacts within the same group, but individuals in different groups make no contacts. For ease of exposition, assume interest lies in quantifying the effect of vaccination which is a one-time event before start of an outbreak. Let *A*_*ij*_ = 1 if individual *j* in each group *i* is vaccinated and *A*_*ij*_ = 0 otherwise.

Let ***A***_***i***_ = (*A*_*i*1,_ *A*_*i*2_, … , *A*_*iN*_) and ***A***_***i*−*j***_ = (*A*_*i*1,_ *A*_*i*2_, … , *A*_*ij***−**1_, *A*_*ij*+1,_ … , *A*_*iN*_), hereafter referred to as allocation programs.^11^ Let **a**_***i***_ and **a**_***i***,**−*j***_ denote possible realizations of ***A***_***i***_ and ***A***_***i***,**−*j***_, respectively. Let 𝒜(*N*) denote the set of all possible 2*^N^* vaccine allocations for a group of size *N*, for which **a**_***i***_ ∈ 𝒜(*N*).

Let *Y*_*ij*_(**a**_***i***_) denote the potential binary outcome for individual *j* in group *i* with allocation program **a**_***i***_ and let *Y*_*ij*_(**a**_***i***,**−*j***_, ***a)*** denote the potential binary outcome when individual *j* has vaccination status *a* and the rest of group *i* has vaccination status **a**_***i***,**−*j***_.

### Individual, group, and population average potential outcomes

Hudgens and Halloran^12^ define *marginal individual average potential outcome* as

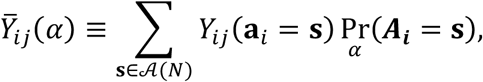

and *individual average potential outcome*^12^ as:

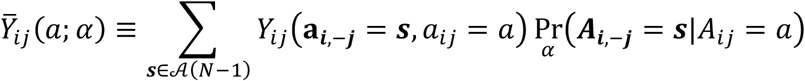

where Pr(⋅) is the probability distribution of vaccine allocation program ***A***_***i***_ with parameter *α* ∈ [0,1] representing the proportion vaccinated within group *i*. Specifically, Pr(⋅) is the probability distribution of ***A***_***i***_ conditional on 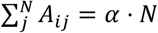. Note here we use type A parameterization, which gives same effect definitions as type B parameterization suggested by VanderWeele and Tchetgen Tchetgen^11^ when *N* is large. Definitions of Pr (⋅), type A, and type B parameterizations are given in eAppendix 1. Hudgens and Halloran^12^ further define (marginal) group average potential outcomes 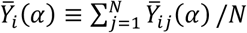 and 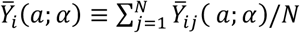 by averaging over individuals within groups. They also define (marginal) population average potential outcomes^12^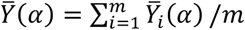 and 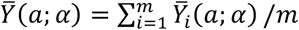

### Population Average Direct, Indirect, Overall, and Total Causal Effects

Hudgens and Halloran define *population average direct casual effect*^12^ as 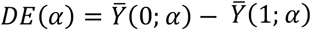, comparing 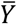 when an individual is unvaccinated versus when vaccinated, holding fixed proportion vaccinated (*α* ). They define *population average indirect casual effect*^12^ as 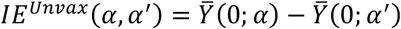, comparing 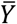 for an unvaccinated individual in a group with *α* proportion vaccinated versus with *α*′, hereafter referred to as indirect effect for the unvaccinated. As suggested previously,^17^ indirect effect can be analogously defined for a vaccinated individual: 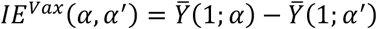, hereafter referred to as indirect effect for the vaccinated. They also define *population average total causal effect*^12^ as 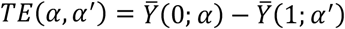, comparing 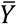 an individual when they are unvaccinated in a group with *α* proportion vaccinated versus when they are vaccinated in a group with *α*′. Following from the definition, *TE*(*α*, *α*′) can be decomposed as 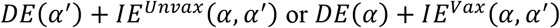. Finally, they define *population average overall casual effect*^12^ as 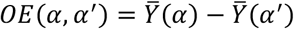, comparing 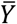 for a typical individual in a group with *α* proportion vaccinated versus with *α*′.

### Overall Effect Partitioning

Previously, Hudgens and Halloran^12^ showed that when *α* = 0, *OE*(*α*, *α*′) is the weighted sum *α*′ ⋅

*TE*(*α*, *α*′) + (1 – *α*′) ⋅ *IE*^*Unvax*^(*α*, *α*′). Here, to establish direct impact as a lower bound on overall impact (Section 3), we generalize *OE*(*α*, *α*′) partitioning to any *α*′ > *α* in Theorem 1.

#### Theorem 1

*(overall effect partitioning)*

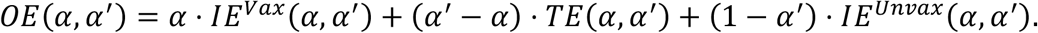

Theorem 1 is proved in eAppendix 2 and graphically illustrated in Figure 1. Theorem 1 expresses *OE*(*α*, *α*′) as a weighted average of three effects: 1) *IE*^*Vax*^(*α*, *α*′) , 2) *TE*(*α*, *α*′) , and 3)*IE*^*Unvax*^(*α*, *α*′). Intuitively, if individuals are classified by vaccination status under a pair of counterfactuals wherein the group has *α* or *α*′ proportion vaccinated (*α*′ > *α*), then *IE*^*Vax*^(*α*, *α*′) is in operation for proportion *α* of individuals who are vaccinated under both counterfactuals; *TE*(*α*, *α*′) is in operation for proportion *α*′ − *α* of individuals who are vaccinated under *α*′ but not *α*; and *IE*^*Unvax*^ (*α*, *α*′) is in operation for proportion 1 − *α*′ of individuals who are unvaccinated under both counterfactuals.

**Figure 1.**
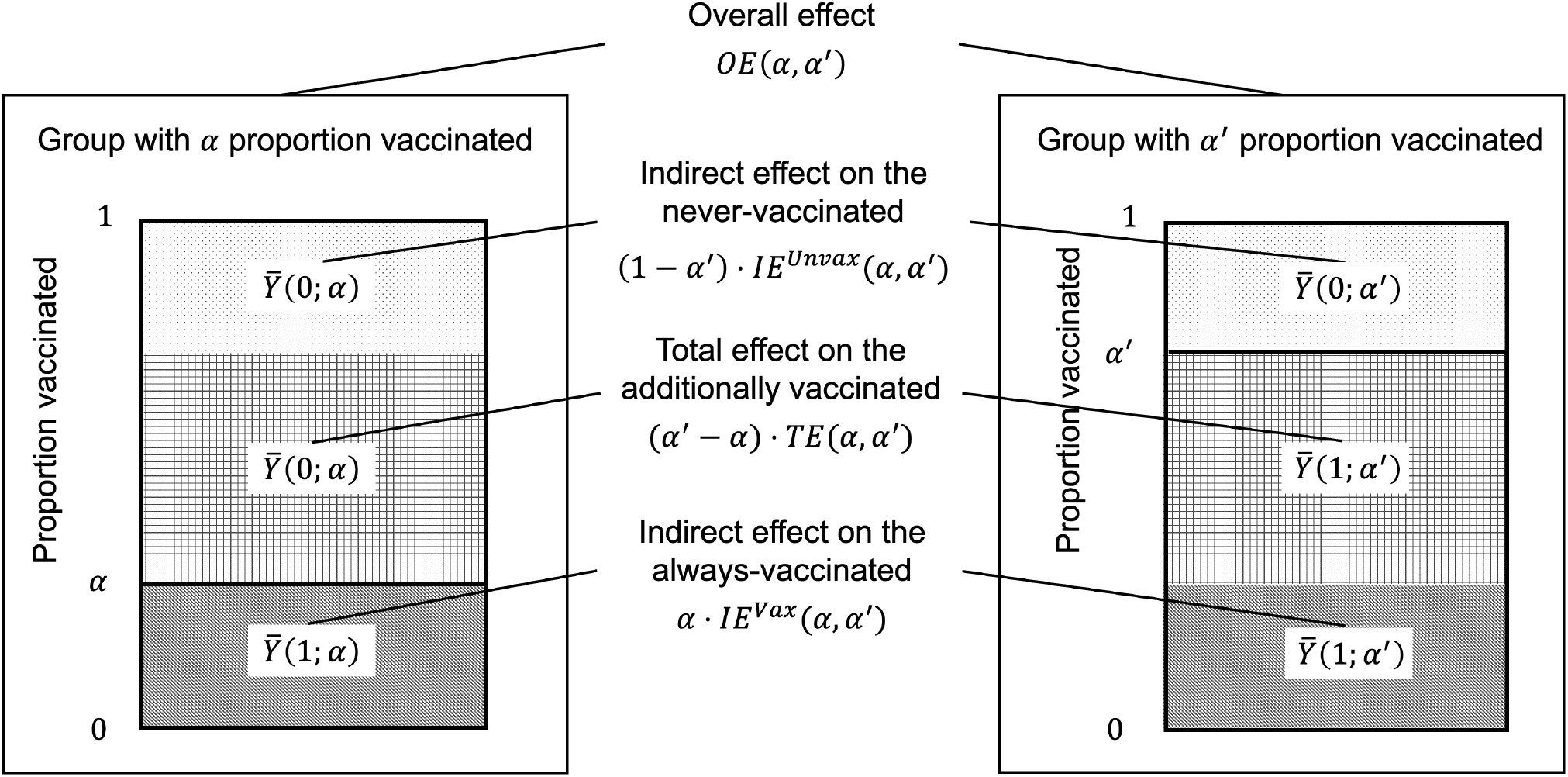
Graphical illustration on partitioning overall effect. The two rectangles represent a pair of counterfactuals wherein the group has *α* or *α*′ proportion vaccinated (*α*′ > *α*). Individuals fall into three categories based on their vaccination status under the counterfactuals: 1) The dotted region represents those who are unvaccinated under both counterfactuals (hereafter referred to as “never-vaccinated”) and for whom *IE*^*Unvax*^(*α*, *α*′) is in operation; 2) the gridded region represents those who are unvaccinated under *α* but vaccinated under *α*′ and for whom *TE*(*α*, *α*′) is in operation (hereafter referred to as “additionally-vaccinated”); and 3) the stripped region represents those who are vaccinated under both counterfactuals (hereafter referred to as “always-vaccinated”) and for whom *IE*^*Vax*^(*α*, *α*′) is in operation. Theorem 1 shows that *OE*(*α*, *α*′) is a weighted average of three effects: 1) *IE*^*Unvax*^(*α*, *α*′), 2) *TE*(*α*, *α*′), and 3) *IE*^*Vax*^(*α*, *α*′), each weighted by the proportion of individuals for whom the effect is in operation respectively: 1) 1 − *α*′ for the never-vaccinated, 2) *α*′ − *α* for the additionally-vaccinated, and 3) *α* for the always-vaccinated.

## 3. ESTIMANDS FOR ANALYSES OF AVERTED AND AVERTIBLE OUTCOMES

Up until now, all effect estimands (except indirect effect for vaccinated)^17^ have been previously defined by Hudgens and Halloran in a two-stage randomized trial. To define estimands for analyses of averted and avertible outcomes, we now expand on the terminology of two-stage randomization. Although a two-stage randomized trial is rarely conducted in practice, our goal is to be explicit about the definition of estimands in recent observational studies on averted and avertible outcomes, and to place these observational studies in the context of target trial emulation, which is playing a growing role in causal inference.

The original overall effect (OE) and direct effect (DE) are defined for individual risk,^10^ and their magnitudes are not comparable because OE and DE use different denominators. However, empirical observational studies of averted and avertible deaths use count outcomes, assuming vaccination prevents more deaths overall than directly (i.e., OE multiplied by total population size is greater than multiplying DE with number of vaccinated individuals in the presence of indirect protection for unvaccinated individuals).^4–9^ To verify this assumption, Section 3 defines two estimands for averted and avertible outcomes—direct impact and overall impact—that align with estimands targeted by literature on vaccine-averted and avertible outcomes.^1–9^ Sections 4 and 5 then verify conditions under which direct impact is or is not a lower bound on overall impact.

This paper adds to existing literature by: (1) Introducing an additional time index *t* to examine outcomes at multiple timepoints post-vaccination, (2) defining direct and overall impact (i.e., corollaries of direct and overall effect estimands for count outcomes) to be used in averted and avertible outcome analyses, (3) defining the relationships between direct and overall impact, and (4) most importantly, using analytical and simulation approaches to identify conditions under which direct impact may or may not be a lower bound on overall impact.

### Notation

To examine direct and overall impact over time, we incorporate time index *t* so that 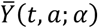 denotes population average cumulative incidence of having developed the outcome by time *t*, and similarly for 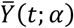 and all the effect estimands in Section 2. *t* = 0 denotes start of follow-up. In principle, *t* can be of any timescale, while *t* is in days in the simulations in Section 5.

Now let *α*_1_ denote a particular vaccination proportion chosen to be implemented in the group, let *α*_2_ denote some hypothetical higher proportion (i.e., *α*_2_ > *α*_1_), and let *α*_0_ denote some hypothetical lower proportion (i.e., *α*_0_ < *α*_1_).

### Motivating Examples and Causal Questions

During the COVID-19 pandemic, determining the total number of infections (or deaths) averted by vaccination has been of great public health interest.^1–7,18–20^ Vaccine-averted infections (or deaths) is an impact estimand based on the causal question: How many infections (or deaths) have been averted under the particular proportion vaccinated (*α*_1_) compared to the counterfactual in the absence of vaccination (*α*_0_ = 0)?

Alternatively, researchers have also estimated the vaccine-avertible deaths—those that could have been averted by vaccination but were not because of a failure to vaccinate.^8,9^ The causal question is: How many infections (or deaths) could have been averted under full vaccination (*α*_2_ = 1), but were not averted given the particular proportion vaccinated (*α*_1_)?

In general, estimands are defined by comparing number of infections (or deaths) in a typical group under the particular proportion vaccinated ( *α*_1_) versus a lower ( *α*_0_ ) or higher ( *α*_2_ ) proportion.^9^ We term these as *impact* estimands because previous literature has referred to vaccine-averted infections (or deaths) as (population) impact of vaccination.^5,7,18,19^

### Overall Impact

For *α*_0_ < *α*_1_, overall impact is defined as:

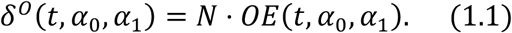

For *α*_2_ > *α*_2_, overall impact is defined as:

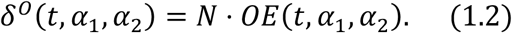

Overall impact directly answers the two aforementioned causal questions on quantifying vaccine-averted and -avertible outcomes. For reasons clarified later, we force overall impact to have the same sign when *α*_1_ is compared to a lower (*α*_0_) or higher (*α*_2_) hypothetical value.

Note mathematical modelling studies implicitly refer to overall impact when estimating vaccine-averted deaths by simulating the epidemic trajectory under a hypothetical proportion vaccinated (e.g., *α*_0_ = 0) and comparing it with the trajectory under the particular vaccination campaign (*α*_1_).

Furthermore, by Theorem 1 and *TE*(*t*, *α*_0_, *α*_0_) = *DE*(*t*, *α*_0_) + *IE*^*Unvax*^ (*t*, *α*_0_, *α*_1_) , we decompose:

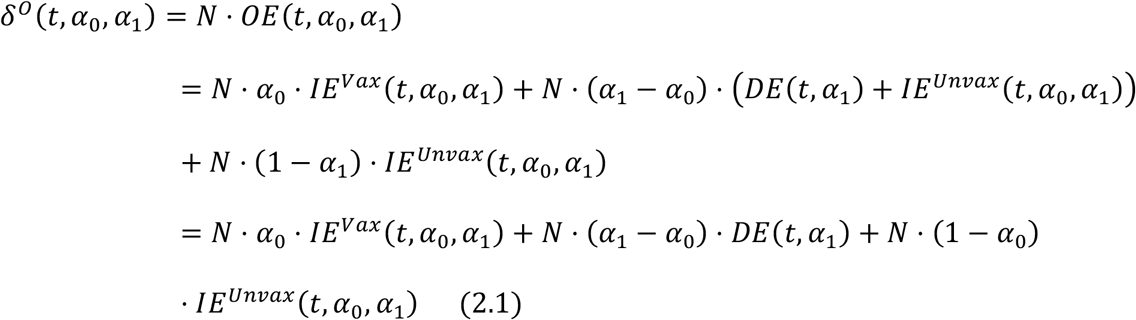

for *α*_1_ > *α*_0_ . The first term on the right-hand side of last line of equation (2.1) scales *IE*^*Vax*^(*t*, *α*_0_, *α*_1_) by number vaccinated under *α*_1_ . The second term scales *DE*(*t*, *α*_1_) by the additional number vaccinated under *α*_1_ compared to *α*_0_. The third term scales *IE*^*Unvax*^(*t*, *α*_0_, *α*_1_) by number unvaccinated under *α*_0_.

Similarly, by Theorem 1 and *TE*(*t*, *α*_1,_ *α*_2_) = *DE*(*t*, *α*_1_) + *IE*^*Vax*^(*t*, *α*_1,_ *α*_2_), we decompose:

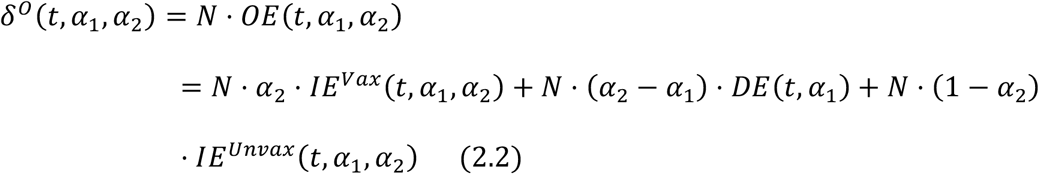

for *α*_2_ > *α*_1_. The first term on the right-hand side of last line of equation (2.2) scales *IE*^*Vax*^(*t*, *α*_0_, *α*_1_) by number vaccinated under *α*_2_ . The second term scales *DE*(*t*, *α*_1_) by the additional number vaccinated under *α*_2_ compared to *α*_1_. The third term scales *IE*^*Unvax*^(*t*, *α*_0_, *α*_1_) by number unvaccinated under *α*_2_.

### Direct Impact

Because overall impact (*δ*^*o*^ ) contrasts two different vaccination proportions, at minimum researchers need to observe two noninteracting groups where each is “assigned” to one of the vaccination strategies to estimate overall impact.^12^ Ideally, vaccination would be assigned via a two-stage randomized trial. However, these trials are rare because they are expensive and hard to justify.^21^ Instead, most empirical studies only observe a single group under one vaccination proportion (*α*_1_), in which case researchers can only estimate an impact corollary of direct effect. Examples include observational studies comparing discrete hazards or incidence rates between vaccinated and unvaccinated individuals based on national vaccine data systems.^4–9^ These studies multiply direct effect with number vaccinated (or unvaccinated) to estimate outcomes averted (or avertible) via direct impact among vaccinated (or unvaccinated) individuals,^4–9^ and then generally assume that it is lower than overall impact in the entire population.^4–6,8^ Here, we formalize this definition of direct impact and show how it relates to overall impact.

Some empirical observational studies have estimated deaths averted via direct impact among vaccinated individuals^4–7^ using formulas similar to *N* ⋅ (*α*_1_ – *α*_0_) ⋅ *DE*(*t*, *α*_1_) by setting *α*_0_ = 0, while others have estimated deaths avertible via direct impact among unvaccinated individuals^8,9^ using formulas similar to *N* ⋅ (*α*_2_ – *α*_1_) ⋅ *DE*(*t*, *α*_1_) by setting *α*_2_ > *α*_1_ (Note these studies have also considered increases in proportion vaccinated over time, such as under a vaccine rollout, which here we ignore for simplicity). Following the literature,^4–9,18^ let direct impact (*δ*^*D*^) for any

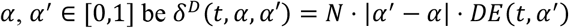

In particular, for *α*_1_ > *α*_0_, we have

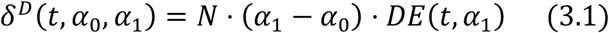

and for *α*_2_ > *α*_1_,

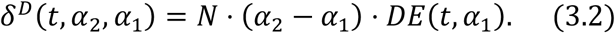

Note *δ*^*D*^(*t*, *α*_0_, *α*_1_) or *δ*^*D*^(*t*, *α*_2_, *α*_1_) is not a meaningful causal estimand by itself because *DE*(*t*, *α*_1_) is conditional on *α*_1_ only and does not account for change in DE when proportion vaccinated is *α*_0_ or *α*_2_ instead of *α*_1_. Importantly, now direct impact can be a lower bound on overall impact. This is because Theorem 1 decomposes *OE* into *TE, IE*^*Unvax*^, and *IE*^*Vax*^, such that we can map direct impact onto *N* ⋅ |*α*′ − *α*| ⋅ *DE*(*t*, *α*′) in equations 2.1 and 2.2, which are now written as:

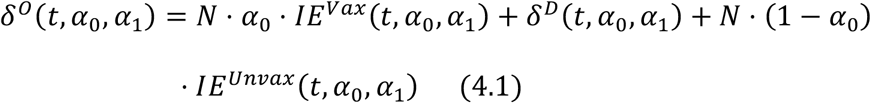

for *α*_1_ > *α*_2_, and

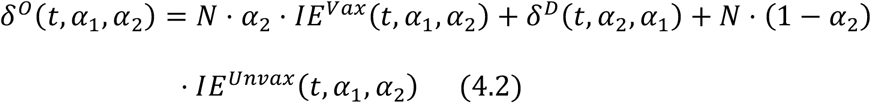

for *α*_2_ > *α*_1_. Since overall impact has the same sign when *α*_1_ is compared to a lower (*α*_0_) or higher (*α*_2_) hypothetical value, we formalize the assumption often made in the empirical vaccine-averted and -avertible outcome studies—that is, direct impact is a lower bound on overall impact by considering Claim 1:

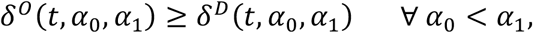

and

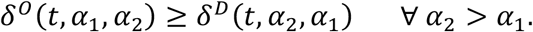

If Claim 1 is true, direct impact, which can be estimated using commonly available data,^4–9,18^ is a lower bound on overall impact that is relevant for policy-making and retrospective policy evaluation requiring samples from a population of groups as in two-stage randomized trials.^12^

Following the literature, we consider two special cases of Claim 1. In Claim 1a, a particular vaccination proportion *α*_1_ is compared to a hypothetical of no vaccination (*α*_0_ = 0) to quantify vaccine-averted outcomes.

Claim 1a (vaccine-averted outcomes):

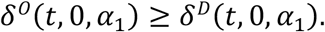

In words, Claim 1a asserts that for *α*_1_ > *α*_0_ = 0, outcomes averted via direct impact of current vaccination among vaccinated individuals is a lower bound on total vaccine-averted outcomes among both vaccinated and unvaccinated individuals.

In Claim 1b, *α*_1_ is compared to a hypothetical of near-universal vaccination (*α*_2_ = 0.9 > *α*_1_) to quantify vaccine-avertible outcomes, following the literature.^9^

Claim 1b (vaccine-avertible outcomes):

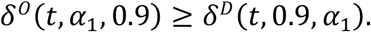

In words, Claim 1b asserts that for *α*_2_ = 0.9 > *α*_1_, outcomes avertible via direct impact of current vaccination among some unvaccinated individuals is a lower bound on vaccine-avertible outcomes among both vaccinated and unvaccinated individuals. Note we do not compare to *α*_2_ = 1 because DE would be undefined under full vaccination, and there would not be an epidemic under full vaccination with a highly effective vaccine.

Based on the partitioning in equations 4.1 and 4.2, Claims 1a and 1b hold if *IE*^*Vax*^ and *IE*^*Unvax*^ are non-negative. However, it is not immediately intuitive when that occurs. Therefore, we describe a transmission model to check Claims 1a and 1b under various scenarios (Section 4), and then describe conditions under which direct impact is or is not a lower bound on overall impact (Section 5).

## 4. TRANSMISSION MODEL

### The SIRD Model with Vaccination at Baseline

An SIRD model represents a well-mixed group in a two-stage randomized trial assuming partial interference.^12^ To simulate direct and overall impact, we simulate a typical group with a large size under a pair of counterfactual vaccination proportions. In this simulation, (marginal) group average potential outcome is equivalent to (marginal) population average potential outcome because there is only one group in the population. The model consists of four states for a vaccinated or unvaccinated individual—susceptible, infectious, recovered, and death due to infection. We assume that the group is randomly assigned to a vaccination policy wherein the proportion to be vaccinated is *α*, and individuals are randomly assigned a vaccination status *a* at baseline (*a* = 1 for vaccinated and *a* = 0 for unvaccinated; for equation [5], subscript *v* denotes vaccinated and *u* for unvaccinated) based on their group proportion. The vaccine is “leaky” in protecting against infection and infection-related death—that is, vaccination reduces susceptibility by *θ* against infection (i.e., vaccine efficacy against infection [*VE*_*infection*_] is (1 − *θ*) • 100%) and reduces susceptibility by *κ* against death (i.e., vaccine efficacy against death given infection [*VE*_*death*|*infection*_] is (1 − *κ*) • 100%). Individuals mix homogeneously such that each vaccinated or unvaccinated susceptible individual is equally likely to contact any infectious individual. Vaccinated and unvaccinated infectious individuals are equally contagious. The transmission dynamics are:

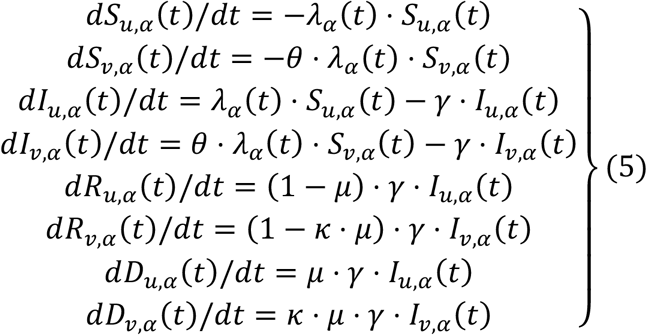

where *γ* = recovery rate, 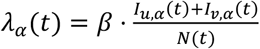 is the hazard rate of infection, with *β* the number of effective contacts made by a typical infectious individual per unit time, and *μ* = probability of death due to infection. In equation (5), *S*_*u,α*_ (*t*) and *S*_*v,α*_ (*t*) denote, respectively, the number of susceptible individuals who are unvaccinated and vaccinated, *I*_*u,α*_ (*t*) and *I*_*v,α*_ (*t*) for the infectious individuals, *R*_*u,α*_ (*t*) and *R*_*v,α*_ (*t*) for the recovered individuals who are no longer at risk, and *D*_*u,α*_ (*t*) and *D*_*v,α*_ (*t*) for those who died due to infection. *N*(*t*) denotes the sum of all compartments at time *t*. eFigure 1 shows the model flowchart, and eTable 1 shows parameter values used in simulation.

### Software

All simulations and visualization are conducted using R 4.2.2 (R Foundation for Statistical Computing, Vienna, Austria).^22^ All models are implemented using R package *odin*.^23^ Code is available at https://github.com/katjia/impact_estimands.

## 5. WHEN IS DIRECT IMPACT A LOWER BOUND ON OVERALL IMPACT?

### Rationale

The SIRD model has many simplifying assumptions compared to real-world settings. However, if one can show a counterexample to Claim 1 based on the simplest SIRD model, then Claim 1 is not guaranteed to be true in more general and realistic models. To identify counterexamples of Claim 1, we consider common violations to assumptions on time-invariant parameters so that indirect effects can be negative: (1) Number of effective contacts may increase over time due to meteorological factors,^24^ lifting of non-pharmaceutical interventions,^25^ and seasonal variation in social contacts, (2) infection-fatality risk may increase over time,^26^ (3) waning immunity clearly occurred in the Delta and Omicron waves of COVID-19 pandemic.^27,28^

### Scenarios

We check the Claims under 5 scenarios (Table). Scenario 1 refers to the SIRD model in equation (5) with time-invariant parameters. eTable 1 lists model parameters for which one or two parameters vary under each Scenario separately: Scenario 2 increases number of effective contacts made by a typical infectious individual per day (*β*) from 0.15 to 0.6 from Day 300 onwards; Scenario 3 increases probability of death due to infection (*μ*) from 0.01 to 0.1 from Day 300 onwards; Scenario 4 allows both *VE*_*infection*_and *VE*_*death*|*infection*_to wane linearly after Day 100 reaching 0% at Day 300; and Scenario 5 combines Scenarios 2 (increasing *β*) and 4 (waning VEs).

**TABLE.**
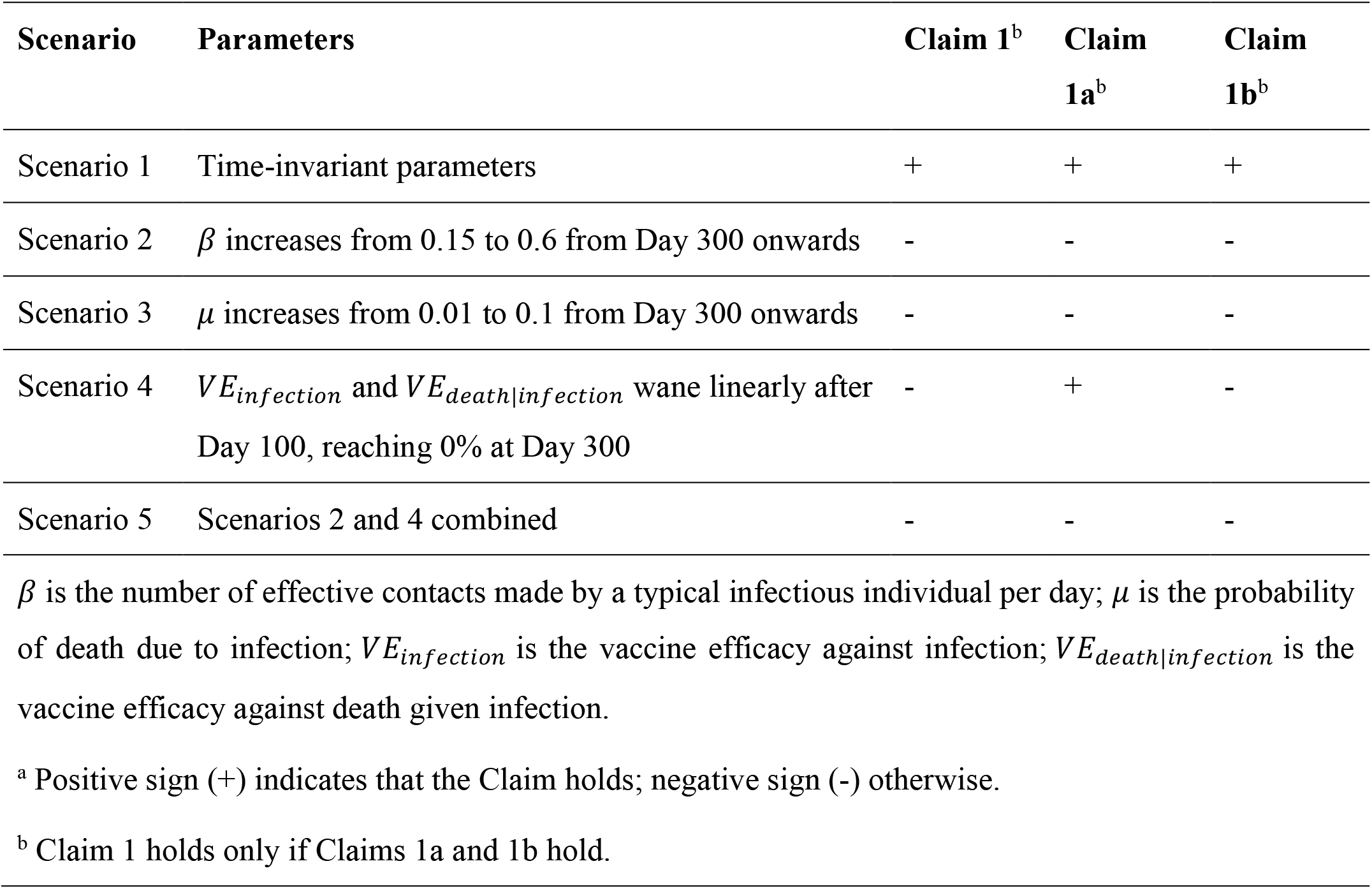
Scenarios under which the Claims may or may not hold ^a^.

### Proof of Claim 1 at the end of outbreak under Scenario 1

For Scenario 1 (i.e., time-invariant parameters), in eAppendix 4, we prove that Claim 1 (i.e., direct impact is a lower bound on overall impact for any two proportions vaccinated) holds in the SIRD model at the end of outbreak (i.e., at *t* → ∞).

### Simulations

For *t* < ∞, we first verify Claims 1a and 1b (special cases of Claim 1) through simulation based on parameters specified in eTable 1 and initial conditions in eTable 2. We specify *α*_0_ = 0 versus *α*_1_ = 0.7 in the pair of trajectories to verify Claim 1a and *α*_1_ = 0.7 versus *α*_2_ = 0.9 to verify Claim 1b. If Claims 1a and 1b both hold for the specified parameters, Latin hypercube sampling is conducted to generate alternative sets of proportions vaccinated and model parameters to verify the full Claim 1. If only one of Claim 1a or 1b holds, Latin hypercube sampling is conducted to verify whether the Claim holds under alternative proportion vaccinated and model parameters (e.g., trying alternative values for *α*_1_ while fixing *α*_0_ = 0 for Claim 1a).

Briefly, Claim 1 only holds under Scenario 1 (i.e., time-invariant parameters), but not under any other Scenarios. The Table summarizes the results. Figures 2 and 3 show trajectories of direct and overall impact throughout the epidemic to verify Claims 1a and 1b, respectively. Figures 2 and 3 show that Claims 1a and 1b hold under Scenario 1. Moreover, Latin hypercube sampling verifies that Claim 1 holds under Scenario 1 (eAppendix 5). Under Scenario 2 where *β* increases, Claims 1a and 1b do not hold: Direct impacts are not lower bounds on overall impacts (Figures 2 and 3) due to negative indirect effects (eFigures 3 to 5). Under Scenario 3 where *μ* increases, Claims 1a and 1b hold for infections but not deaths due to negative indirect effects for death (eFigures 3 to 5). Under Scenario 4 where VEs wane, only Claim 1a (vaccine-averted outcomes) holds (Figure 2). Latin hypercube sampling verifies that Claim 1a holds under Scenario 4 (eAppendix 5). However, Claim 1b (vaccine-avertible outcomes) does not hold (Figure 3) due to negative indirect effects (eFigures 4 and 5). Finally, under Scenario 5 where *β* increases and VEs wane, Claims 1a and 1b do not hold.

**FIGURE 2.**
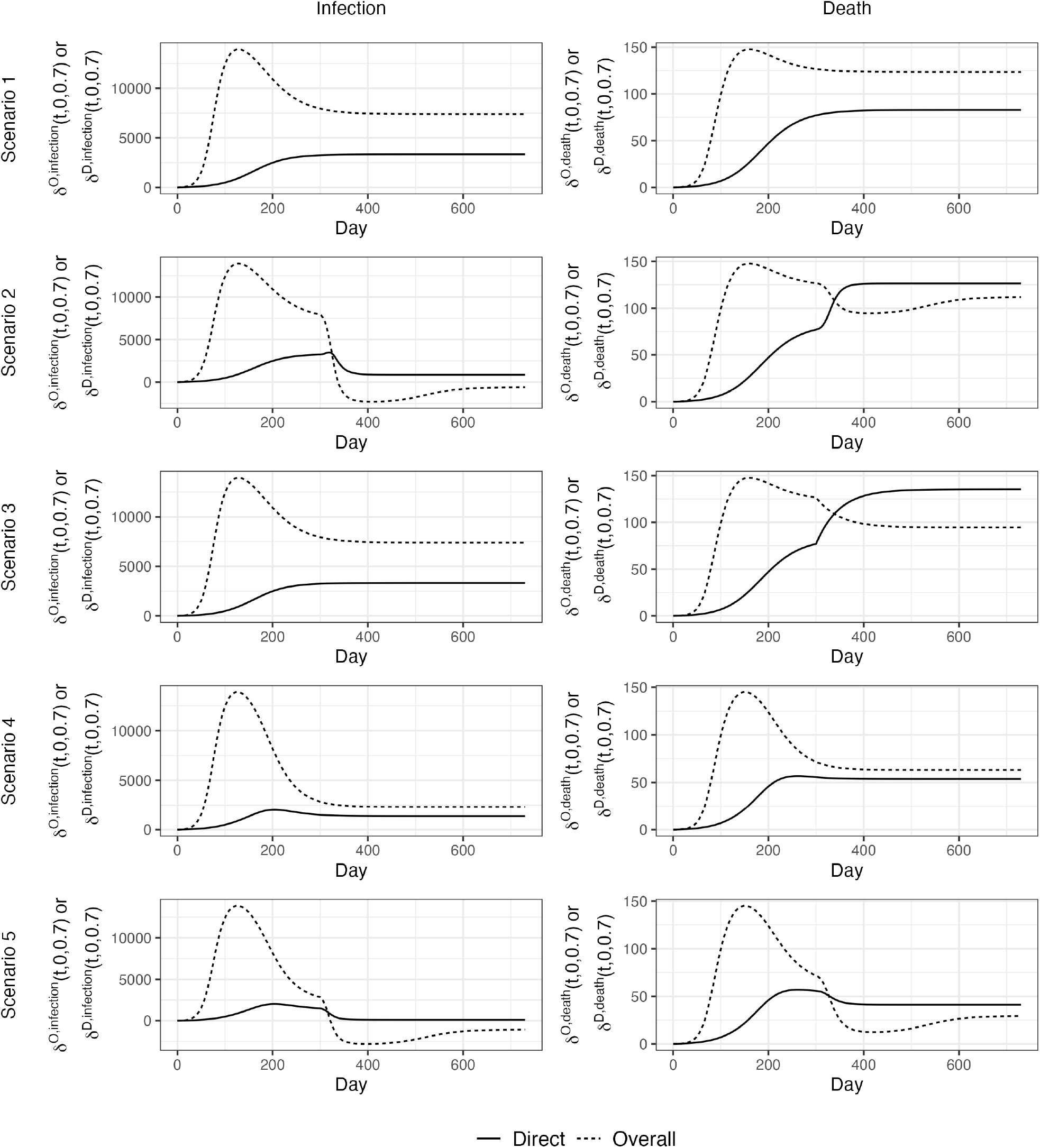
Direct impact and overall impact given **α**_0_ = 0 and **α**_1_ = 0. 7 to verify Claim 1a. Scenario 1, all parameters are time-invariant; Scenario 2, the number of effective contacts made by a typical infectious individual per day (*β*) increases from 0.15 to 0.6 at Day 300; Scenario 3, probability of infection-related death (*μ*) increases from 0.01 to 0.1 at Day 300; Scenario 4, vaccine efficacies against infection and death start to wane linearly after Day 100 reaching 0% at Day 300; and Scenario 5, the combination of Scenarios 2 and 4.

**FIGURE 3.**
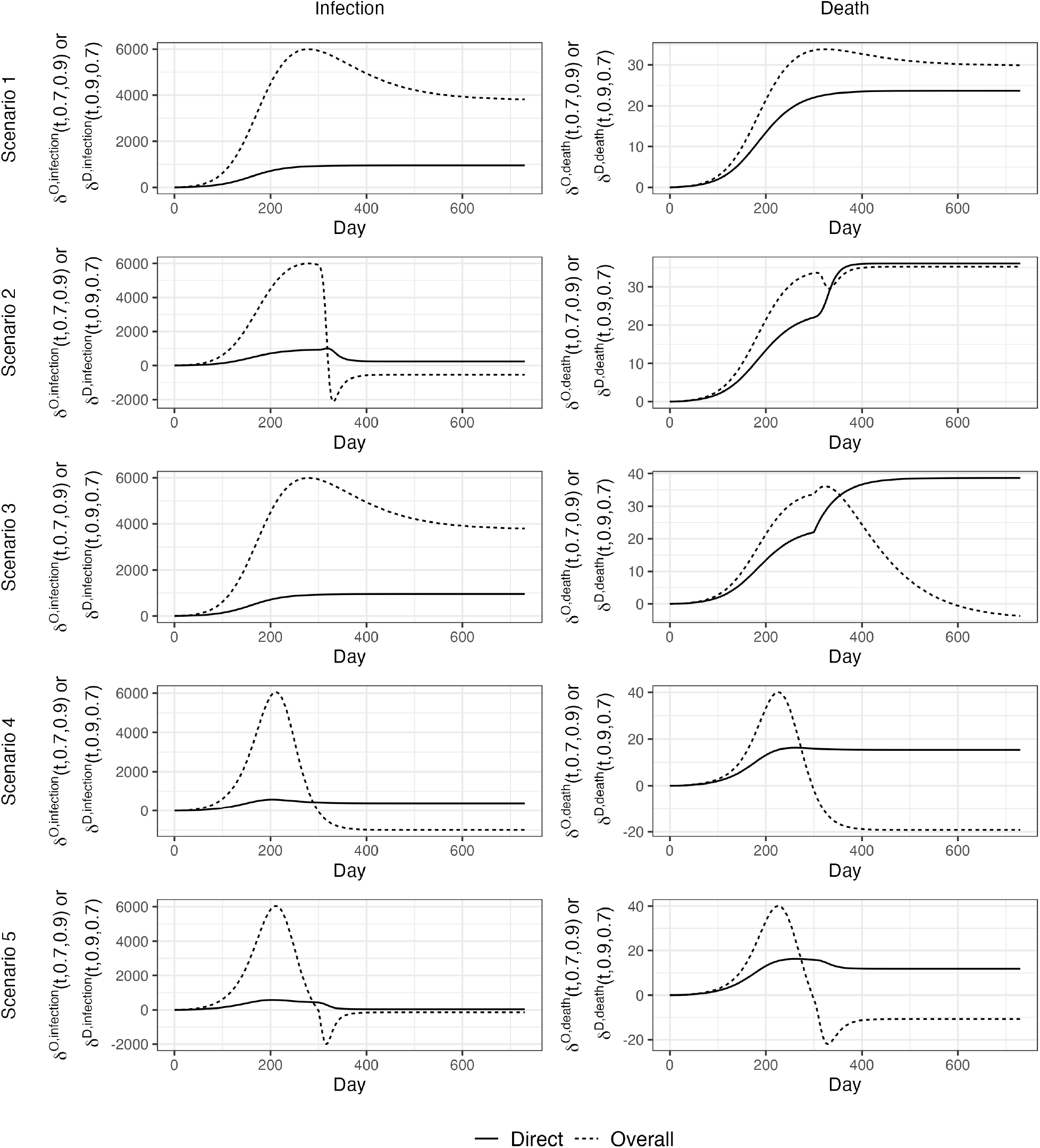
Direct impact and overall impact given **α**_1_ = 0. 7 and **α**_2_ = 0. 9 to verify Claim 1b. Scenario 1, all parameters are time-invariant; Scenario 2, the number of effective contacts made by a typical infectious individual per day (*β*) increases from 0.15 to 0.6 at Day 300; Scenario 3, probability of infection-related death (*μ*) increases from 0.01 to 0.1 at Day 300; Scenario 4, vaccine efficacies against infection and death start to wane linearly after Day 100 reaching 0% at Day 300; and Scenario 5, the combination of Scenarios 2 and 4.

## 6. DISCUSSION

Motivated by recent research on deaths averted by COVID-19 vaccination, this study adjusts estimands defined by Hudgens and Halloran to accommodate analyses on averted and avertible outcomes due to infectious disease interventions, thereby enabling researchers to distinguish the estimands when conducting and interpreting related studies. An epidemic model is simulated to verify the commonly held claim that outcomes averted (or avertible) via direct impact among the vaccinated (or unvaccinated) individuals is a lower bound on overall impact. Based on the SIRD model, the lower bound fails when transmission or fatality parameter increases, or vaccine efficacies wane, implying that the lower bound is not guaranteed to hold for more general and realistic models. Consequently, it would be overly optimistic for empirical studies^4–9^ to assume that they have estimated a lower bound on true number of averted (or avertible) outcomes through estimating the direct impact without examining the directions of indirect effects.

When indirect effects for the vaccinated and unvaccinated are both positive (that is, higher vaccination coverage yields positive number of averted or avertible outcomes), direct impact is a lower bound on overall impact (equations 4.1 and 4.2). Subsequently, we show in eAppendix 4 that, at the end of epidemic, deaths averted via direct impact would be a lower bound on overall impact in a SIRD model given time-invariant parameters and vaccination at baseline, thanks to the positive indirect effects, a finding proven more recently in a preprint by Lin et al. (2024).^29^ In general, indirect effects may be positive because vaccination reduces the infectious individuals at a given time, such that infection-naïve individuals are less likely to be infected.^30^ However, as shown above, there are scenarios under which direct impact is not a lower bound on overall impact due to negative indirect effects (Scenarios 2–5).

First, when the number of effective contacts made by a typical infectious individual per day (*β*) increases over time (Scenario 2), overall impact on infection decreases (and can be negative) because at the early stage of outbreak, the less-vaccinated group has many infected and recovered with sterilizing immunity; while the more-vaccinated group has more susceptible (i.e., infection-naïve) individuals who have escaped earlier infections and will experience a higher force of infection at later time (eFigure 6). *β* is affected by meteorological factors,^24^ behavioral factors (e.g., usage of personal protective equipment),^31^ biological factors (e.g., changes in host immunity, evolution of strains),^32^ non-pharmaceutical interventions, and seasonal variation in social contacts.

Second, when probability of infection-related death (*μ*) increases over time (Scenario 3), overall impact on death decreases because the extensively vaccinated group(s) has more who escape earlier infections and then experience higher fatality at later stage of outbreak (eFigure 6). It is plausible for lethality of pathogens to increase over time: Disease severity increased in the autumn wave of the 1918 flu pandemic compared to the spring-summer wave of the same pathogen in that year.^26^ Increasing lethality implies that vaccination at beginning of outbreak can decrease overall impact by postponing cases. On the other hand, if fatality rate increases with infection peak due to the sudden shortage of healthcare resources, overall impact on death will be more positive because vaccines delay infection and flatten the epidemic curve. Likewise, if infection-fatality rates decline progressively due to improvements in care,^33,34^ then vaccination that delays the epidemic can have amplified positive overall impact.

Third, overall impact may become negative when vaccine efficacies wane (Scenario 4). In particular, vaccination proportion of *α*_2_ = 0.9 may result in more infections and deaths than the proportion of *α*_1_ = 0.7. eAppendix 8 discusses this scenario in greater detail.

In addition, if there were multiple risk groups with heterogenous susceptibility to adverse outcomes and heterogeneous mixing patterns, vaccination for a subgroup could cause negative indirect effect in other subgroups by increasing risk for more severe complications. For example, empirical evidence showed that low rubella vaccination coverage in children increased rubella incidence in the 15-and-over and incidence of congenital rubella in newborns.^35^

The current study has some limitations. First, vaccination is assumed to be a one-time event at baseline before start of outbreak, but in reality, vaccine rollout is continuous over time and may occur during outbreak. Second, throughout, we have considered the case where vaccination occurs at random. However, in most empirical settings there may be strong confounding due to staged rollout of vaccines and differences in vaccine acceptance by behavioral and health characteristics. Such confounding if uncontrolled threatens the validity of inferences about the effects, whether or not a bound is valid in ideal (unconfounded) circumstances.

Another limitation is that in the scenarios considered, changes in lethality and transmission were assumed to occur at a fixed time, whereas in reality they might well occur either in response to pathogen evolution^36^ or to behavioral changes that are affected by the epidemic trajectory. However, our goal was not to describe the details of a particular epidemic, but to describe qualitatively conditions under which infectious disease interventions may not prevent more cases overall than directly. Finally, the SIRD model does not consider deaths due to other factors, meaning that simulations are applicable to studies whose outcome of interest is a consequence of infection. Over a short time frame (e.g., 1 year), it is acceptable to consider deaths due to infection only, when other causes of deaths are negligible. We also did not consider possible adverse events after vaccination, although adverse events have important policy implications. Our focus here is impact of interventions on averting disease outcomes. Future studies can extend the estimands to investigate adverse events.

In conclusion, this study examines the commonly held assumption in empirical vaccine-averted and avertible analyses that direct impact is a lower bound on overall impact due to indirect protection. We show that the use of direct impact as a lower bound on overall impact is reliable only under very strong and often unrealistic assumptions. Therefore, it is inadvisable for empirical studies to assume this relation without examining the directions of indirect effects. Alternatively, if researchers want to estimate the averted and avertible outcomes, a transmission model should be used to capture the overall impact, thereby improving clarity about what is estimated despite the expense of making additional modelling assumptions.

## Data Availability

This is a simulation study with code available at https://github.com/katjia/impact_estimands.

## eAppendix of

### eAppendix 1. Type A versus type B parameterizations

In the main text, the potential outcomes 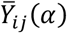 and 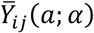 are defined under type A parameterization. eAppendix 1 will define type A and B parameterizations. We assume a large group size *N* such that results are equivalent under both types of parameterizations.^1^ Let *ρ*(***A***_***i***_ = **a**_**i**_; *α*) denote the probability that group *i* receive allocation program **a**_**i**_ given parameter *α*.

#### Definition 1 (A)

According to VanderWeele and Tchetgen Tchetgen,^1^ a parameterization is said to be of type A with parameter *α* = (*N, K*) for group *i* if the allocation program ***A***_***i***_ is randomly allocated conditional on 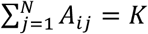 with probability mass function:

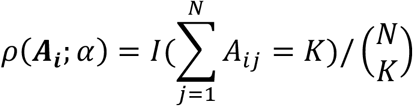

**(B)** A parameterization is said to be of type B with parameter *α* if the allocation program ***A***_***i***_ is randomly assigned to individuals in group *i* according to the known Bernoulli probability mass function:

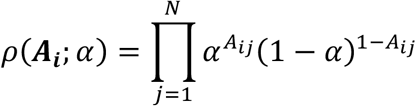

According to VanderWeele and Tchetgen Tchetgen,^1^ Hudgens and Halloran ^2^ defined causal effects under Type A parameterization.

#### Definition 2

The *marginal individual average potential outcome* of Hudgens and Halloran ^2^ is:

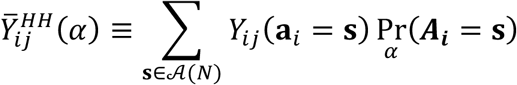

where Pr (***A***_***i***_ = ***s***) = *ρ*(***A***_***i***_ = ***s***; *α*) and *ρ*(***A***_***i***_ = ***s***; *α*) follows Definition 1 (A) under type A parameterization. The *individual average potential outcome* ^2^ is:

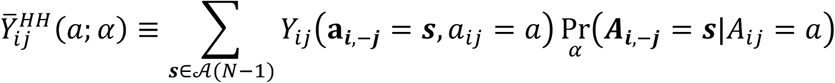

where 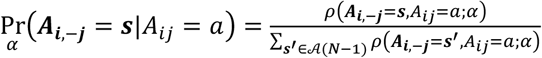 and 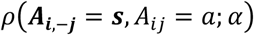 follows

Definition 1 (A) under type A parameterization.

Alternatively, VanderWeele and Tchetgen Tchetgen^1^ proposed the definitions of potential outcomes under Type B parameterization.

#### Definition 3

Under type B parameterization, the *marginal individual average potential outcome*^1^ is:

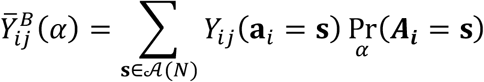

where 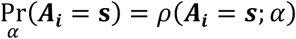 and 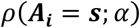 follows Definition 1 (B) under type B parameterization. The *individual average potential outcome*^1^ is:

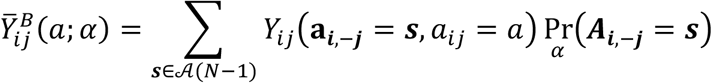

where 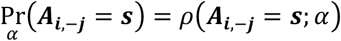 and 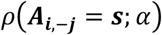 follows Definition 1(B) under type B parameterization.

### eAppendix 2. Proof of Theorem 1 (overall effect partitioning)

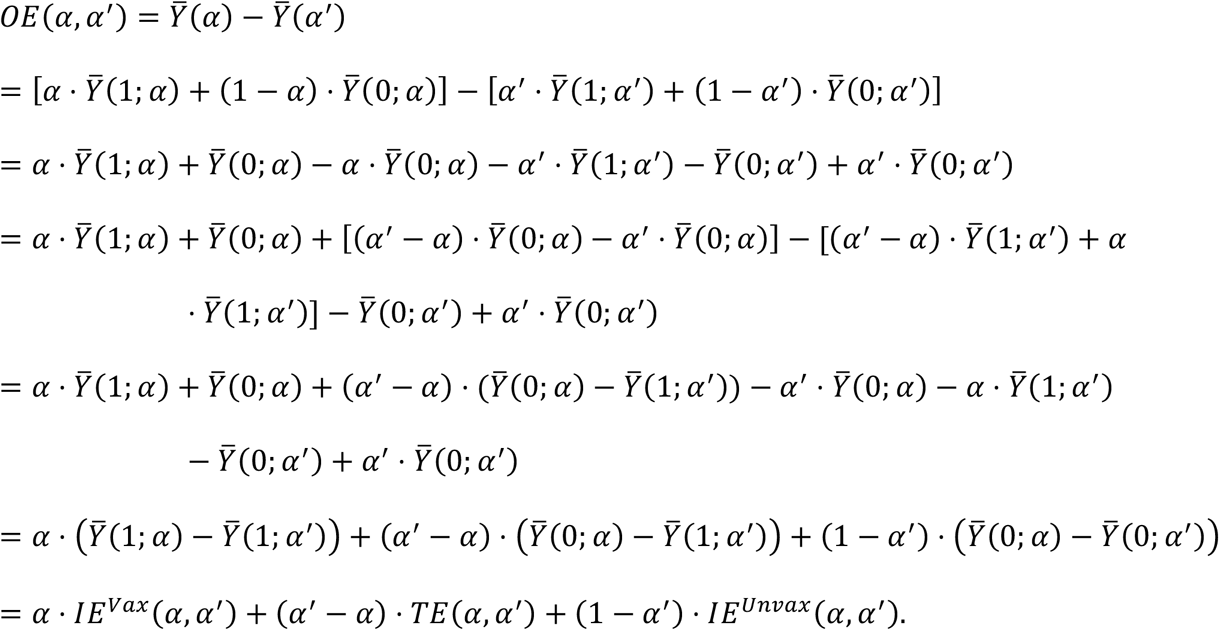

The first line follows by definition; the second follows from line 1 of proof of Theorem 3 from VanderWeele and Tchetgen Tchetgen^1^; the third distributes terms, the fourth adds and subtracts 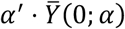 and 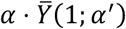; the fifth and sixth rearrange terms; and the last applies definitions of *IE*^*Vax*^, *TE*, and *IE* ^*Unvax*^. The proof can also be shown visually as illustrated in Figure 1.

### eAppendix 3. The susceptible-infected-recovered-death (SIRD) model

#### 1. Model structure

eFigure 1 shows the flowchart for the transmission model described in equation (5) of the main text.

**eFigure 1.**
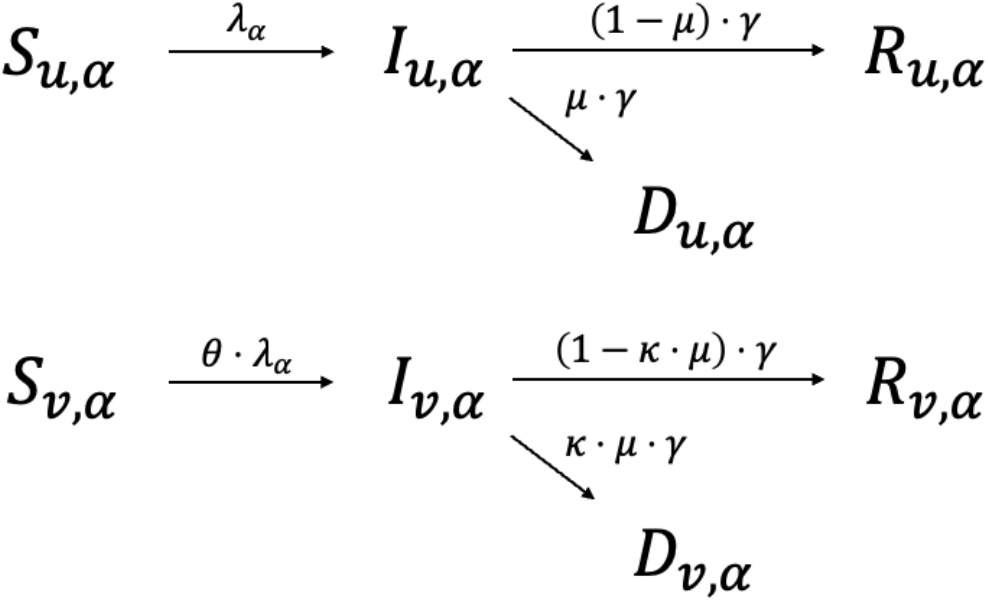
Model flowchart.

#### 2. Model parameters

To verify Claim 1a, a pair of trajectories are simulated with same parameters except for the proportions vaccinated (*α*_0_ = 0 versus *α*_1_ = 0.7). Similarly, to verify Claim 1b, another pair of trajectories are simulated with *α*_1_ = 0.7 versus *α*_2_ = 0.9. Epidemic trajectories are simulated based on the SIRD model in equation (5)—first under Scenario 1 (i.e., time-invariant parameters) and then Scenarios 2 to 5 (i.e., time-dependent parameters). eTable 1 specifies parameter values for simulations. All models are implemented using R package *odin*,^3^ whereas the actual solution of the differential equations is conducted with the *deSolve* package using numerical solvers “lsoda” and “ode45.”^4^

**eTable 1.**
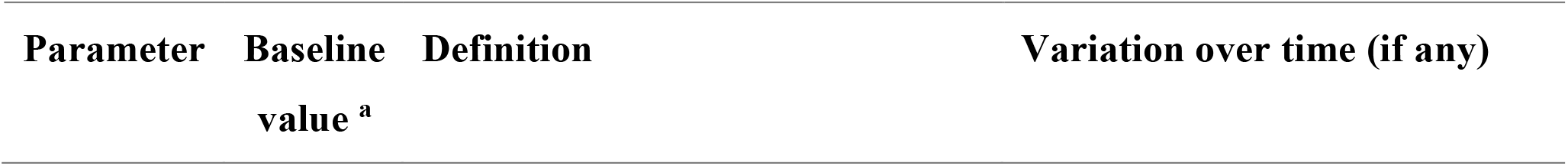
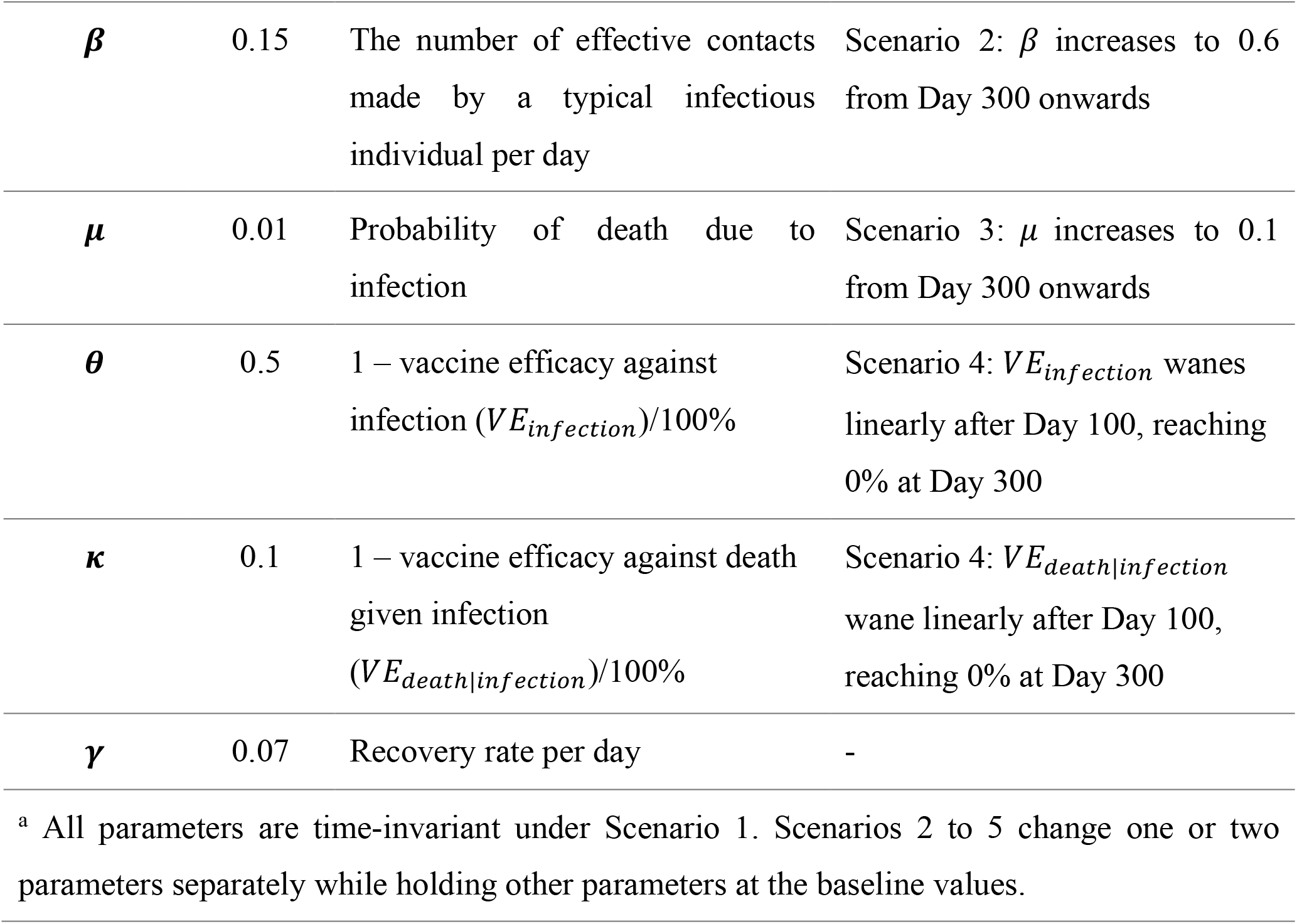
List of parameters for simulations.

**eTable 2.** List of initial conditions in a group wherein the proportion vaccinated is *α*.

| Variable | Initial condition(s) | Definition |
| --- | --- | --- |
| $N(0)$ | 20,000 | Number alive at baseline |
| $S_{u,\alpha}(0)$ | $(20,000 - 20) * (1 - \alpha)$ | Number unvaccinated susceptible at baseline |
| $S_{v,\alpha}(0)$ | $(20,000 - 20) * \alpha$ | Number vaccinated susceptible at baseline |
| $I_{u,\alpha}(0)$ | $20 * (1 - \alpha)$ | Number unvaccinated infectious at baseline |
| $I_{v,\alpha}(0)$ | $20 * \alpha$ | Number vaccinated infectious at baseline |
| $R_{u,\alpha}(0)$ | 0 | Number unvaccinated recovered at baseline |
| $R_{v,\alpha}(0)$ | 0 | Number vaccinated recovered at baseline |
| $D_{u,\alpha}(\mathbf{0})$ | 0 | Number unvaccinated who died due to infection at baseline |
| $D_{v,\alpha}(\mathbf{0})$ | 0 | Number vaccinated who died due to infection at baseline |

### eAppendix 4. Proving Claim 1 under Scenario 1 in the SIRD model at *t* → ∞

#### 1. Definitions

Let 0 ≤ *α*_0_ < *α*_1_ < *α*_2_ ≤ 1. Recall Claim 1:

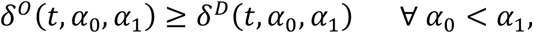

and

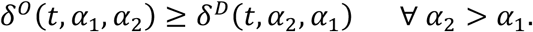

By expanding equations (4.1) and (4.2) and suppressing the time notation (*t*), we have:

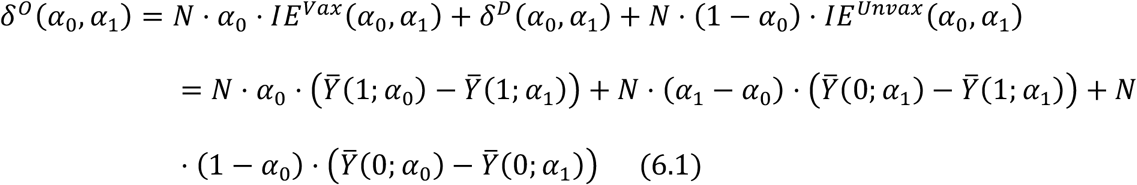

for *α*_1_ > *α*_0_, and

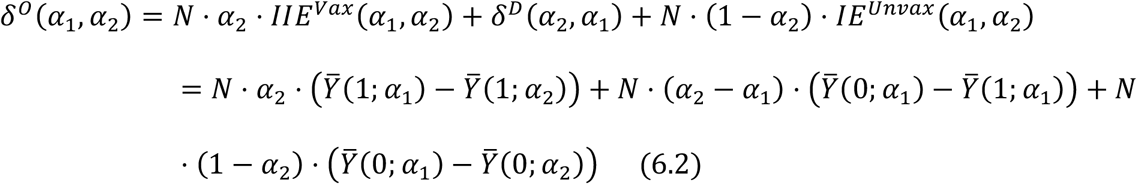

for *α*_2_ > *α*_1_.

We first give an intuitive explanation of *δ*^*D*^(*α*_0_, *α*_1_) being a lower bound of *δ*^*O*^(*α*_0_, *α*_1_). Recall that individuals fall into three categories based on vaccination status under the counterfactuals with *α*_0_ or *α*_1_ proportion vaccinated: 1) Those who are unvaccinated under both counterfactuals (referred to as “never-vaccinated” and represented by the dotted region in Figure 1); 2) those who are unvaccinated under *α*_0_ but vaccinated under *α*_1_ (referred to as “additionally-vaccinated” and represented by the gridded region in Figure 1); and 3) those who are vaccinated under both counterfactuals (referred to as “always-vaccinated” and represented by the stripped region in Figure 1). If we update risk of the “always-vaccinated” under *α*_0_ from 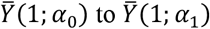 and update risk of the “never-vaccinated” under *α*_0_ from 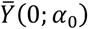 to 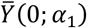, then we have an updated right-hand side of equation (6.1):

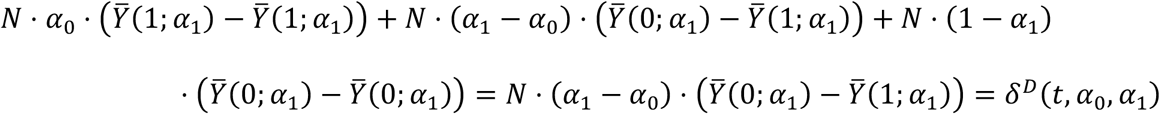

which is reduced to *δ*^*D*^(*α*_0_, *α*_1_) and is lower than the actual overall effect because the original risk 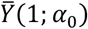 in the first term of the last line of equation (6.1) is higher than the updated 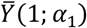 (to be proved in Section 3), and the original risk 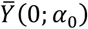 in the last term of the last line of equation (6.1) is also higher than the updated 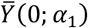 (to be proved in Section 3). Therefore, *δ*^*D*^(*α*_0_, *α*_1_) in the updated equation is lower than *δ*^*O*^(*α*_0_, *α*_1_) in the original equation (6.1) and Claim 1 holds for *α*_1_ > *α*_0_.

Similarly, for *α*_2_ > *α*_1_, if we update risk of “always-vaccinated” under *α*_2_ from 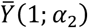 to 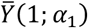 and update risk of “never-vaccinated” under *α*_2_ from 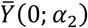 to 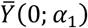, we have on the right-hand side of equation (6.2):

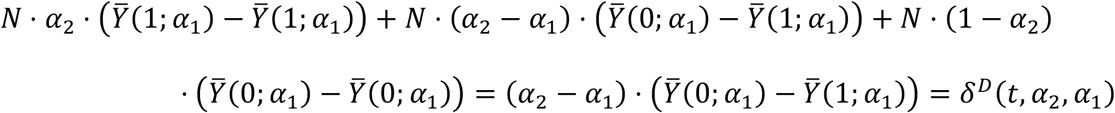

which is reduced to *δ*^*D*^(*α*_2_, *α*_1_) and is lower than the actual overall effect because the original risk 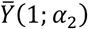 in the first term of the last line of equation (6.2) is lower than the updated 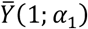 (to be proved in Section 3), and the original risk 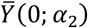 in the last term is also lower than the updated 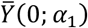 (to be proved in Section 3). Therefore, *δ*^*D*^(*α*_2_, *α*_1_) in the updated equation is lower than *δ*^*O*^(*α*_1,_ *α*_2_) in the original equation (6.2) and Claim 1 holds for *α*_2_ > *α*_1_. Next, we will formally prove Claim 1 under Scenario 1 at *t* → ∞.

#### 2. Sufficient Conditions for Claim 1 to hold

Based on equation (4.1), Claim 1 holds for all *α*_0_ < *α*_1_ when *IE*^*Vax*^ (*t*, *α*_B_, *α*_’_) and *IE*^*Unvax*^(*t*, *α*_0_, *α*_1_) are both non-negative. Similarly, based on equation (4.2), Claim 1 holds for all *α*_2_ > *α*_1_ when *IE*^*Vax*^ (*t*, *α*_1,_ *α*_2_) and *IE*^*Unvax*^ (*t*, *α*_1,_ *α*_2_) are both non-negative.

### 3. Proof of Claim 1 under Scenario 1 in the Susceptible-infected-recovered (SIR) Model at

***t*** → ∞

When probability of death due to infection is zero, the SIRD model in equation (5) reduces to a SIR model stratified by vaccination status. *IE*^*Unvax*^ (∞, *α*_0_, *α*_1_) is positive for *α*_1_ > *α*_0_ in the SIR model by proving that 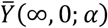, the final epidemic fraction in the unvaccinated individuals (i.e., the fraction of the unvaccinated individuals that became infected at the end of the outbreak), decreases with *α*. Similarly, *IE*^*Vax*^ (∞, *α*_0_, *α*_1_) is positive by proving that 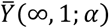, the final epidemic fraction in the vaccinated individuals, also decreases with *α*.

#### 3.1 Proof that 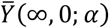 and 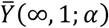 decrease with **α**

To prove that 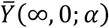 and 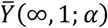 decrease with the proportion vaccinated *α* , we prove that 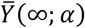, the average final epidemic fraction, decreases with *α*. 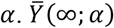 is the weighted average of 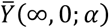 and 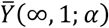.^5^ Assuming that 1) the initial proportion susceptible is close to one, 2) the total group size *N* is near infinite, and 3) transmission events follow a Poisson process, the average final epidemic size is:

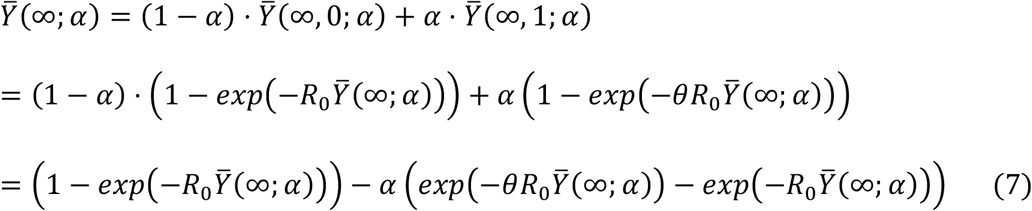

where *θ* = 1 − *VE*_*infection*_/100% given a leaky vaccine, and *R*_0_ is the basic reproduction number, defined as the expected number of new infections caused by a single infected individual during the infectious period in a completely susceptible population. Lin et al. (2024) arrived at the same final epidemic size equation by also considering a leaky vaccine in a stratified susceptible-infected-recovered model. See equation S20 of Lin et al.^6^

Define two separate functions with independent arguments, *Z* and *α* (Miller^5^ used a similar type of argument to arrive at the final epidemic size):

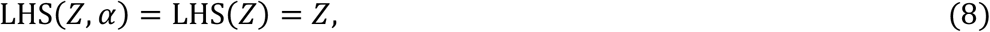

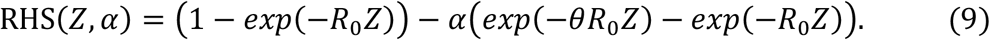

Here the equality LHS(⋅) = RHS(⋅) is not assumed; in particular, RHS(⋅) takes two independent arguments, *Z* and *α*. Plot LHS(*Z*) and RHS(*Z*, *α*) in one graph, where *R*_0_ = 4, *θ* = 0.2, and *α* = 0, 0.3 and 0.7.

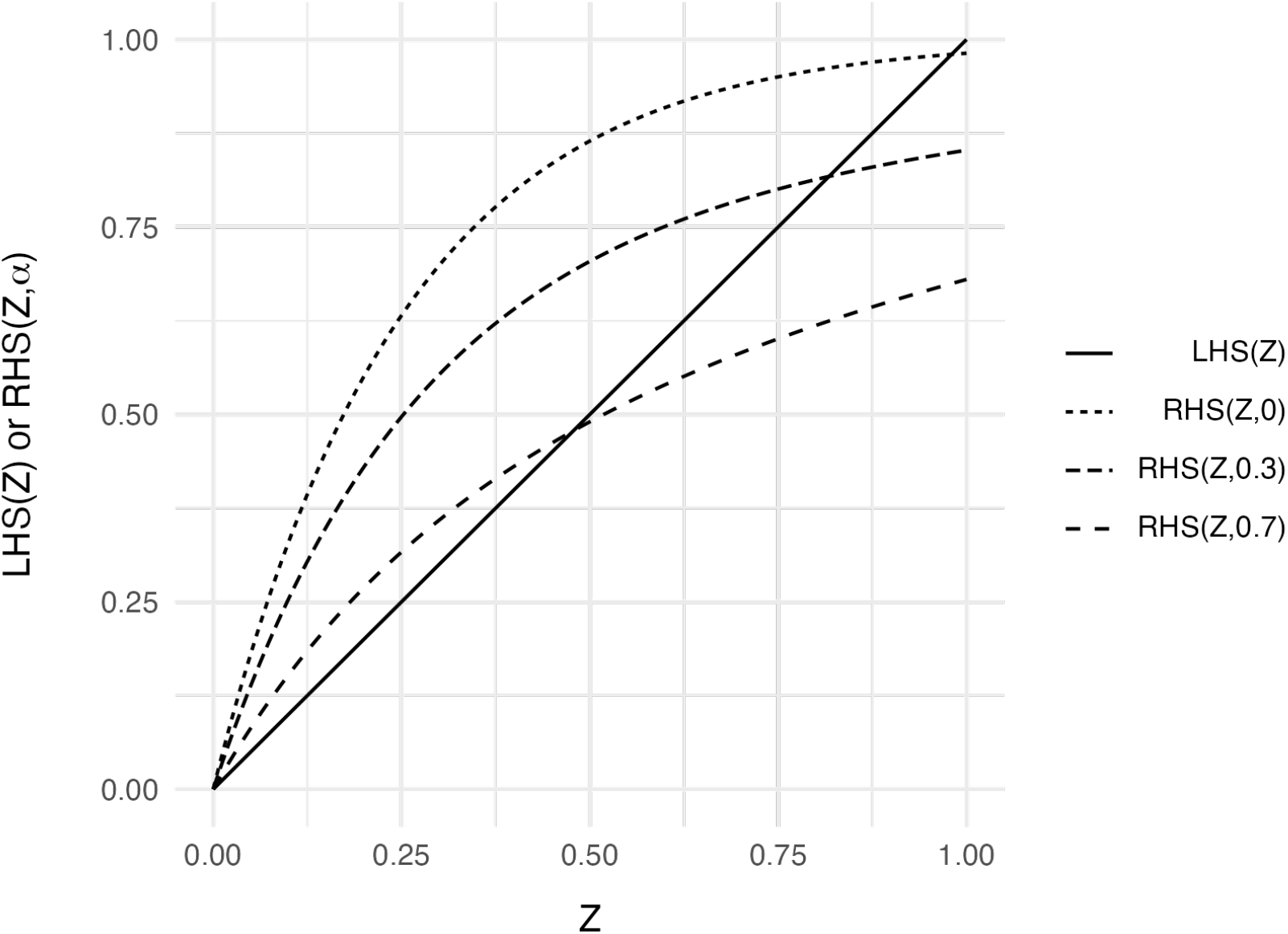

LHS(*Z*) = RHS(*Z*, *α*) when *Z* = 0, which is a trivial solution. We will now prove that if a positive (non-trivial) root exists such that LHS(*Z*) = RHS(*Z*, *α*), the value of this root decreases when *α* increases.

We first show that the first derivative of RHS(*Z*, *α*) w.r.t *Z* at zero is greater than 1, which is the derivative of LHS(*Z*). That is:

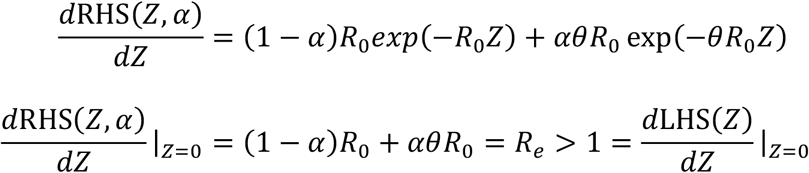

where *R*_*e*_ is the effective reproduction number when proportion *α* of the group is vaccinated with a leaky vaccine.^7^ The above inequality holds because *R*_*e*_ must be greater than 1 in order for any epidemic to occur, meaning that for some *ϵ* > 0, RHS(*ϵ*, *α*) > *LHS*(*ϵ*). In other words, RHS(*Z*, *α*) exceeds LHS(*Z*) for small values of *Z*.

Next, we prove that RHS(*Z*, *α*) is a concave function by computing the second derivative w.r.t *Z*:

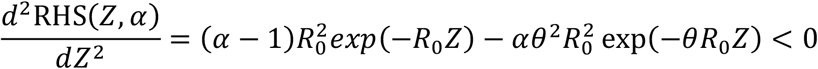

for *α* ∈ [0,1]. Because the second derivative is negative for *α* ∈ [0,1], RHS(*Z*, *α*) is a concave function of *Z* for any possible values of *α*. Taken together, the first and second derivatives indicate that RHS(*Z*, *α*) is above LHS(*Z*) when *Z* is greater than but close to zero, and RHS(*Z*, *α*) bends towards LHS(*Z*) (i.e., the diagonal) and by assumption (that there is a nontrivial root of LHS(*Z*) = RHS(*Z*, *α*)) coincides with the diagonal at some value of *Z* (See plot above).

Finally, consider two values *α*_’_ > *α*_B_ giving rise to two curves RHS(*Z*, *α* = *α*_0_) and RHS(*Z*, *α* = *α*_1_) . Let *α*_1_ = 0.7 and *α*_0_ = 0.3 as the plot illustrates. Because 0 < *θ* < 1 , [*exp*(−*θR*_0_*Z*) − *exp*(−*R*_0_*Z*)] in equation (9) is always positive at any given *Z* > 0. Therefore, RHS(*Z*, *α* = *α*_0_) > RHS(*Z*, *α* = *α*_1_) at any given *Z* > 0 when *α*_1_ > *α*_0_. Suppose the upper curve RHS(*Z*, *α*_0_) meets LHS(*Z*) at *Z* = *z*_0_ . Then RHS(*z*_0_, *α*_’_) < *z*_0_ = LHS(*z*_0_). Given that all expressions are continuous and that RHS(*ϵ*, *α*_*B*_) > LHS(*ϵ*), there must have been some value *z*_1_ to the left of *z*_0_ where RHS(*z*_1,_ *α*_1_) crossed LHS(*z*_1_). In other words, RHS(*z*_1,_ *α*_1_) = LHS(*z*_1_) for some value *z*_1_ < *z*_0_, proving that the root *z*_1_ of RHS(*z*_1,_ *α*_1_) = LHS(*z*_1_) is less than the root *z*_0_ of RHS(*z*_0_, *α*_0_) = LHS(*z*_0_) for *α*_1_ > *α*_0_, QED. These non-trivial roots are unique because RHS(*Z*, *α*) is concave.

Any value *Z* that satisfies LHS(*Z*) = RHS(*Z*, *α*) will satisfy equation (7). Suppose *z*_0_ is a positive root for LHS(*Z*) = RHS(*Z*, *α* = *α*_0_) such that 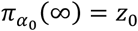, and similarly suppose *z*_1_ is a positive root for LHS(*Z*) = RHS(*Z*, *α* = *α*_1_) such that 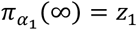. Therefore, we have 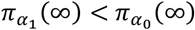 when *α*_1_ > *α*_0_.

Given that 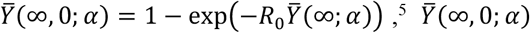 decreases with *α* because 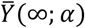decreases with *α* . Similarly, given that 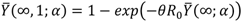 ,^5^ 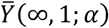 decreases with *α*.

#### 3.2 Proof of *IE*^*unvax*^(∞, **α**_0_, **α**_1_) > 0 and *IE*^*vax*^(∞, **α**_0_, **α**_1_) > 0 for **α**_1_ > **α**_0_

In Section 3.1, we proved that 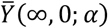 decreases with *α*. Therefore, 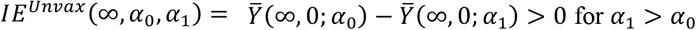 for *α*_1_ > *α*_0_. We also proved that 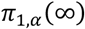decreases with *α*, and therefore 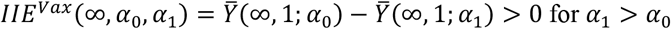 for *α*_1_ > *α*_0_.

Since *IE*^*Unvax*^ (∞, *α*_0_, *α*_1_) and *IE*^*Vax*^ (∞, *α*_0_, *α*_1_) are positive, Claim 1 holds for any *α*_1_ > *α*_0_ at *t* → ∞. The proof in Section 3 also applies to any *α*_2_ > *α*_1_ in showing that *IIE*^*Unvax*^ (∞, *α*_1,_ *α*_2_) and *IIE*^*Vax*^ (∞, *α*_1,_ *α*_2_) are positive, such that Claim 1 holds under Scenario 1 in the Susceptible-infected-recovered (SIR) Model at *t* → ∞.

### 4. Proof of Claim 1 under Scenario 1 in the Susceptible-infected-recovered-death (SIRD)

**Model at *t*** → ∞

Now consider the SIRD model with non-zero probability of death due to infection. Based on the transmission dynamics in equation (5), those who died of infection are fixed proportion of those who became infected at the end of epidemic. We have:

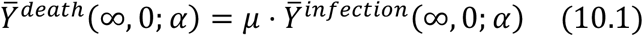

for the unvaccinated individuals and

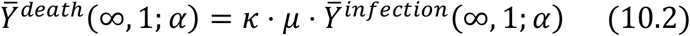

for the vaccinated individuals.

For the unvaccinated individuals, by substituting the right-hand side of equation (10.1) into the equation of *IE*^*Unvax, death*^, we have:

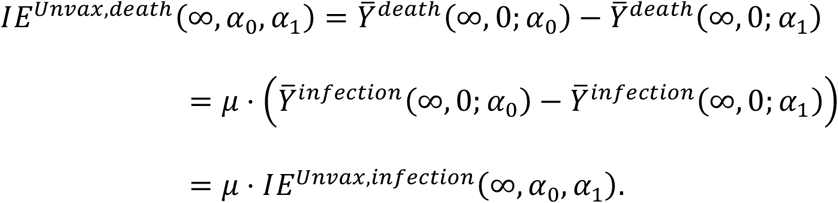

We show that *IE*^*Unvax, death*^ (∞, *α*_0_, *α*_1_) has the same direction as *IE*^*Unvax*, *infection*^ (∞, *α*_0_, *α*_1_) . If *IE*^*Unvax*, *infection*^ (∞, *α*_B_, *α*_’_) is positive (Section 3), so is *IE* ^Unvax, *death*^ (∞, *α*_0_, *α*_1_).

Similarly, for the vaccinated individuals, by substituting the right-hand side of equation (10.2) into the equation of *IE* ^Unvax, death^, we have:

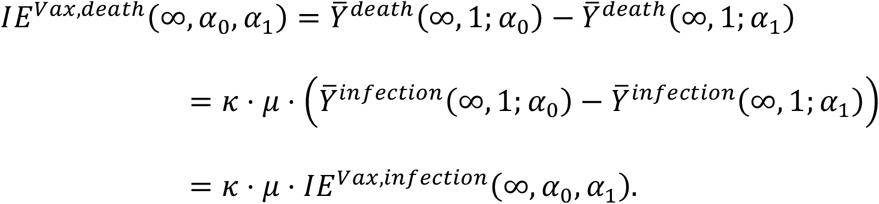

If *IE* ^vax, *infection*^ (∞, *α*_0_, *α*_1_) is positive (Section 3), so is *IE* ^vax, death^ (∞, *α*_0_, *α*_1_).

In sum, *IE* ^vax^ (∞, *α*_B_, *α*_’_) and *IE* ^Unvax^ (∞, *α*_B_, *α*_’_) are positive for both death and infection given *α*_1_ > *α*_0_ (similarly, *IE* ^vax^ (∞, *α*_1,_ *α*_2_) and *IE* ^Unvax^ (∞, *α*_1,_ *α*_2_) are positive for *α*_2_ > *α*_1_). Therefore, Claim 1 holds under Scenario 1 in the Susceptible-infected-recovered-death (SIRD) model at *t* → ∞.

### eAppendix 5. Latin hypercube sampling to verify Claim 1 under Scenario 1 and Claim 1a under Scenario 4

#### 1. Verifying Claim 1 under Scenario 1

As Figures 2 and 3 show, Claims 1a and 1b hold under Scenario 1 given the particular proportions vaccinated (*α*_0_ = 0 versus *α*_1_ = 0.7 for Claim 1a and *α*_1_ = 0.7 versus *α*_2_ = 0.9 for Claim 1b) and parameter values specified in eTable 1. To test the robustness of Claim 1 under alternative parameter combinations, Latin hypercube sampling (LHS) is used to randomly draw 1,000 sets of proportions vaccinated and parameters. Initial conditions are same as in eTable 2. LHS samples are first generated from uniform distributions using the *lhs* package^8^ and then scaled to match the range of each parameter as shown in eTable 3.

**eTable 3.**
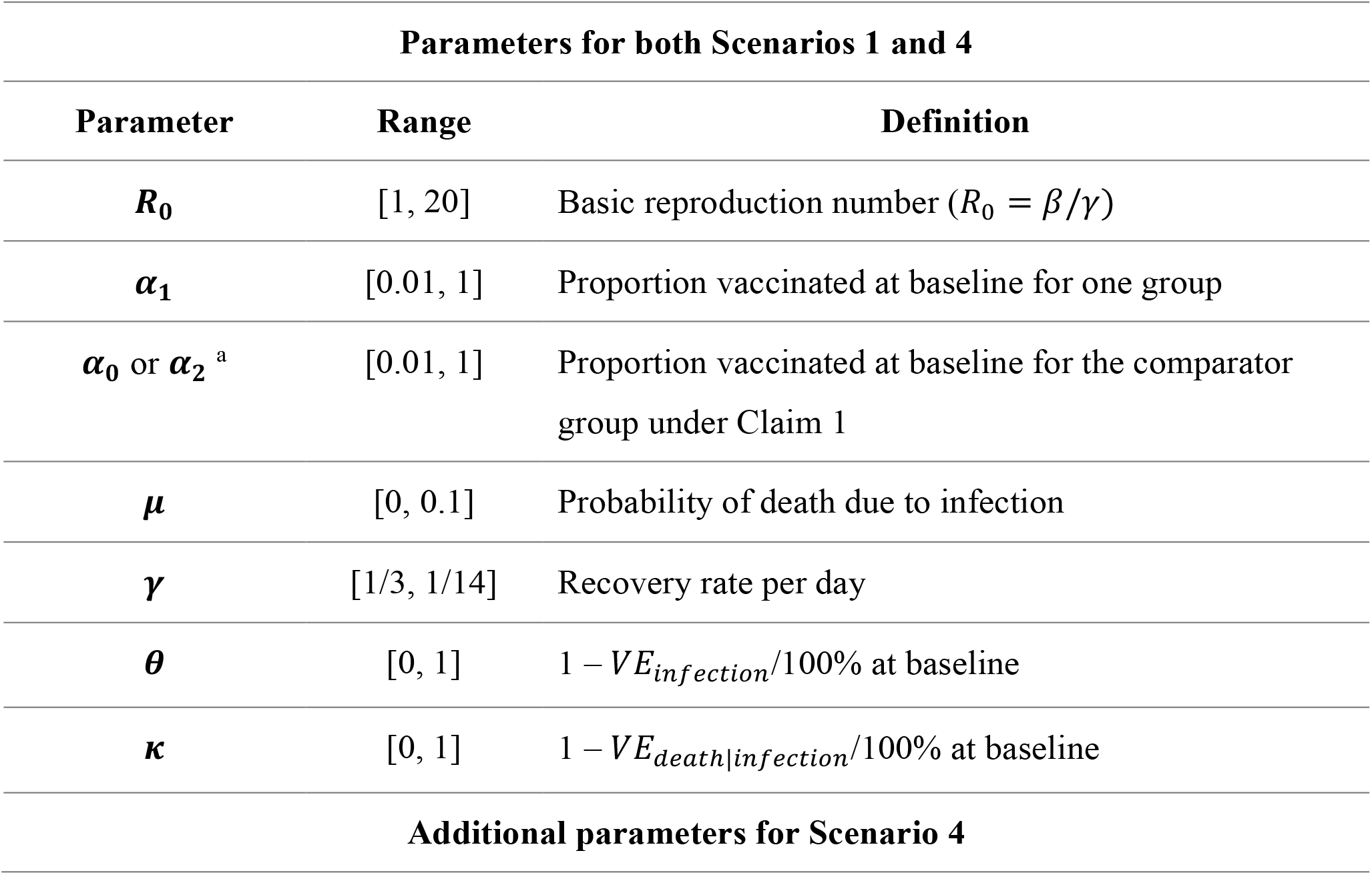
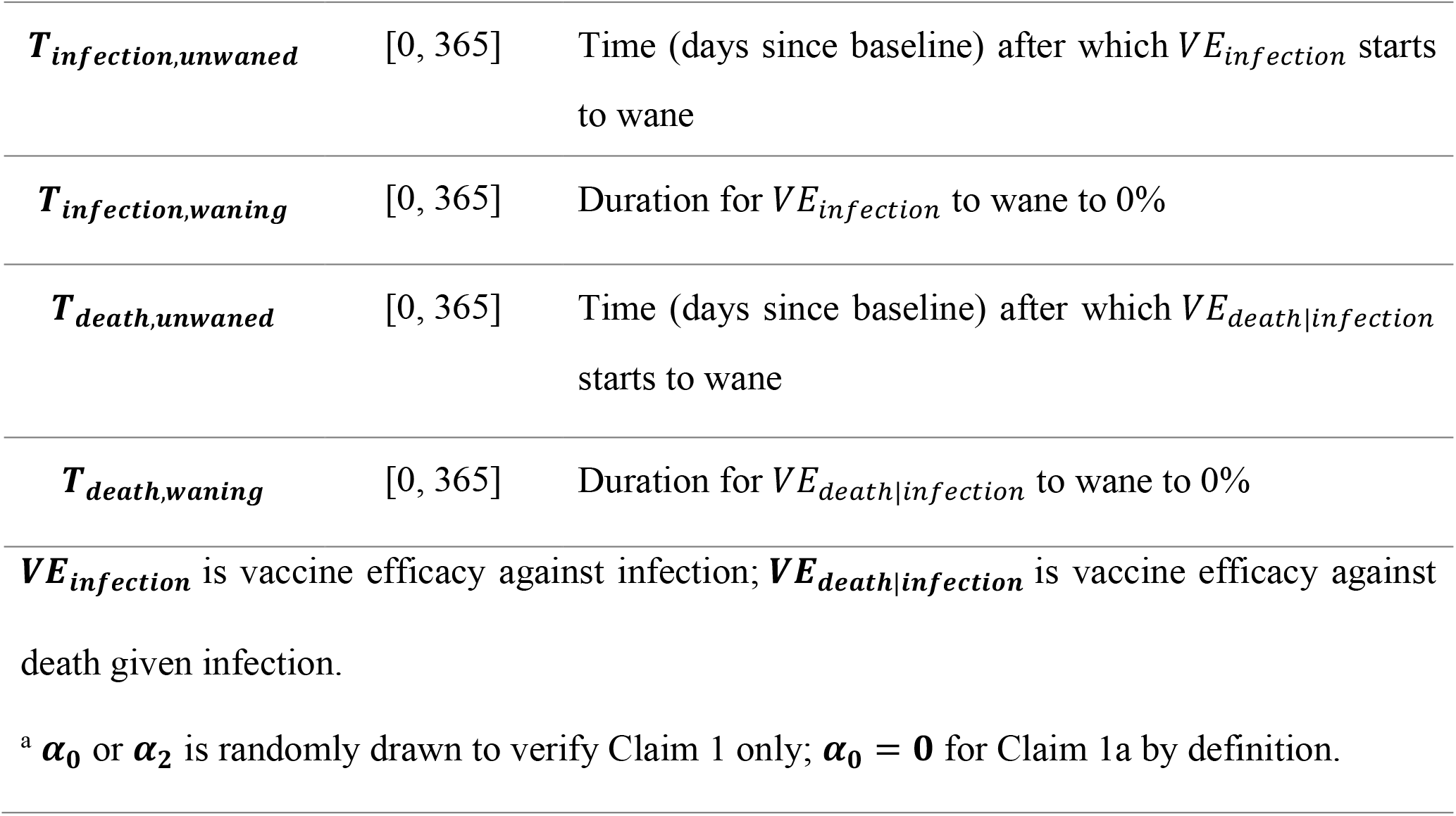
Parameter ranges for Latin hypercube sampling under Scenarios 1 and 4.

| Parameters for both Scenarios 1 and 4 |  |  |
| --- | --- | --- |
| Parameter | Range | Definition |
| $R_0$ | [1, 20] | Basic reproduction number ($R_0 = \beta/\gamma$) |
| $\alpha_1$ | [0.01, 1] | Proportion vaccinated at baseline for one group |
| $\alpha_0$ or $\alpha_2$<sup>a</sup> | [0.01, 1] | Proportion vaccinated at baseline for the comparator group under Claim 1 |
| $\mu$ | [0, 0.1] | Probability of death due to infection |
| $\gamma$ | [1/3, 1/14] | Recovery rate per day |
| $\theta$ | [0, 1] | $1 - VE_{infection}/100\%$ at baseline |
| $\kappa$ | [0, 1] | $1 - VE_{death\vert infection}/100\%$ at baseline |
| Additional parameters for Scenario 4 |  |  |
| $T_{infection,unwaned}$ | [0, 365] | Time (days since baseline) after which $VE_{infection}$ starts to wane |
| $T_{infection,waning}$ | [0, 365] | Duration for $VE_{infection}$ to wane to 0% |
| $T_{death,unwaned}$ | [0, 365] | Time (days since baseline) after which $VE_{death\vert infection}$ starts to wane |
| $T_{death,waning}$ | [0, 365] | Duration for $VE_{death\vert infection}$ to wane to 0% |
| $VE_{infection}$ is vaccine efficacy against infection; $VE_{death\vert infection}$ is vaccine efficacy against death given infection. | | |
| <sup>a</sup> $\alpha_0$ or $\alpha_2$ is randomly drawn to verify Claim 1 only; $\alpha_0 = \mathbf{0}$ for Claim 1a by definition. | | |

To exclude results due to roundoff errors during numerical integration of the differential equations, Claim 1 is disproven based on the criteria *δ*^*o*^ – *δ*^*D*^ < −10^**−**7^ at any time *t*, rather than the conventional criteria of < 0. From the LHS procedure, Claim 1 holds under Scenario 1 for all the 1,000 iterations. eFigure 2 shows direct impact and overall impact for 50 random LHS samples.

#### 2. Verifying Claim 1a under Scenario 4

Figure 2 also shows that Claim 1a holds under Scenario 4 given the parameter values specified in eTable 1. Similarly, LHS is conducted to verify if Claim 1a holds under Scenario 4 with different values for *α*_1_ (while holding constant *α*_0_ = 0) and alternative sets of model parameters; parameter ranges are specified in eTable 3. From the LHS procedure, Claim 1a holds under Scenario 4 for all the 1,000 iterations (eFigure 2).

**eFigure 2.**
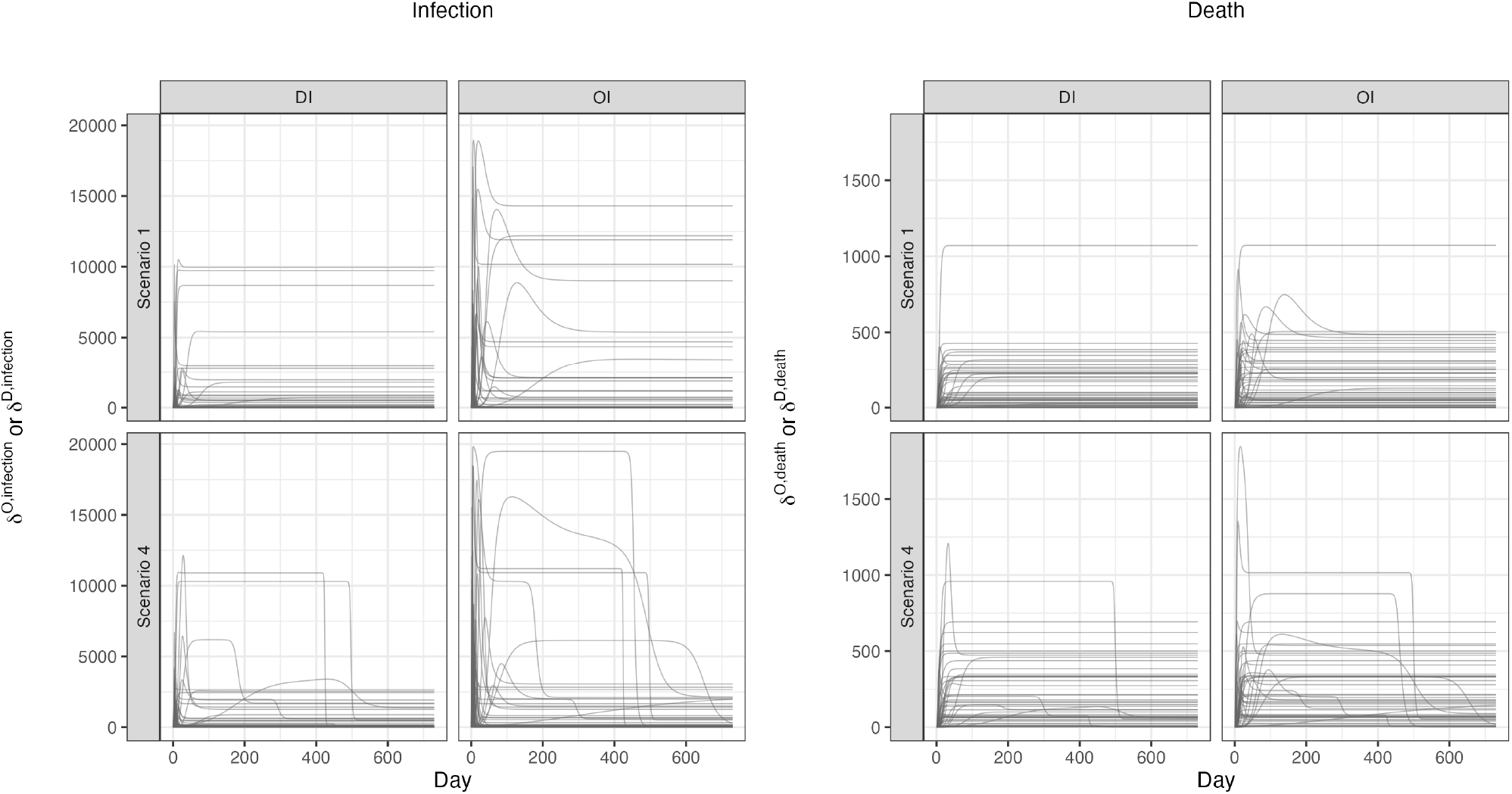
Direct impact and overall impact to verify Claim 1 under Scenario 1 and to verify Claim 1a under Scenario 4 from 50 Latin hypercube samples.

### eAppendix 6. Trajectories of indirect effects for the vaccinated and unvaccinated

Based on equation (4.1), Claim 1a does not hold when *IE* ^Unvax^ (*t*, 0, 0.7) is negative, as shown in eFigure 3. Similarly, based on equation (4.2), Claim 1b may not hold when *IE* ^Unvax^ (*t*, 0.7,0.9) or *IE* ^vax^ (*t*, 0.7,0.9) is negative, as shown in eFigures 4 and 5, respectively.

**eFigure 3.**
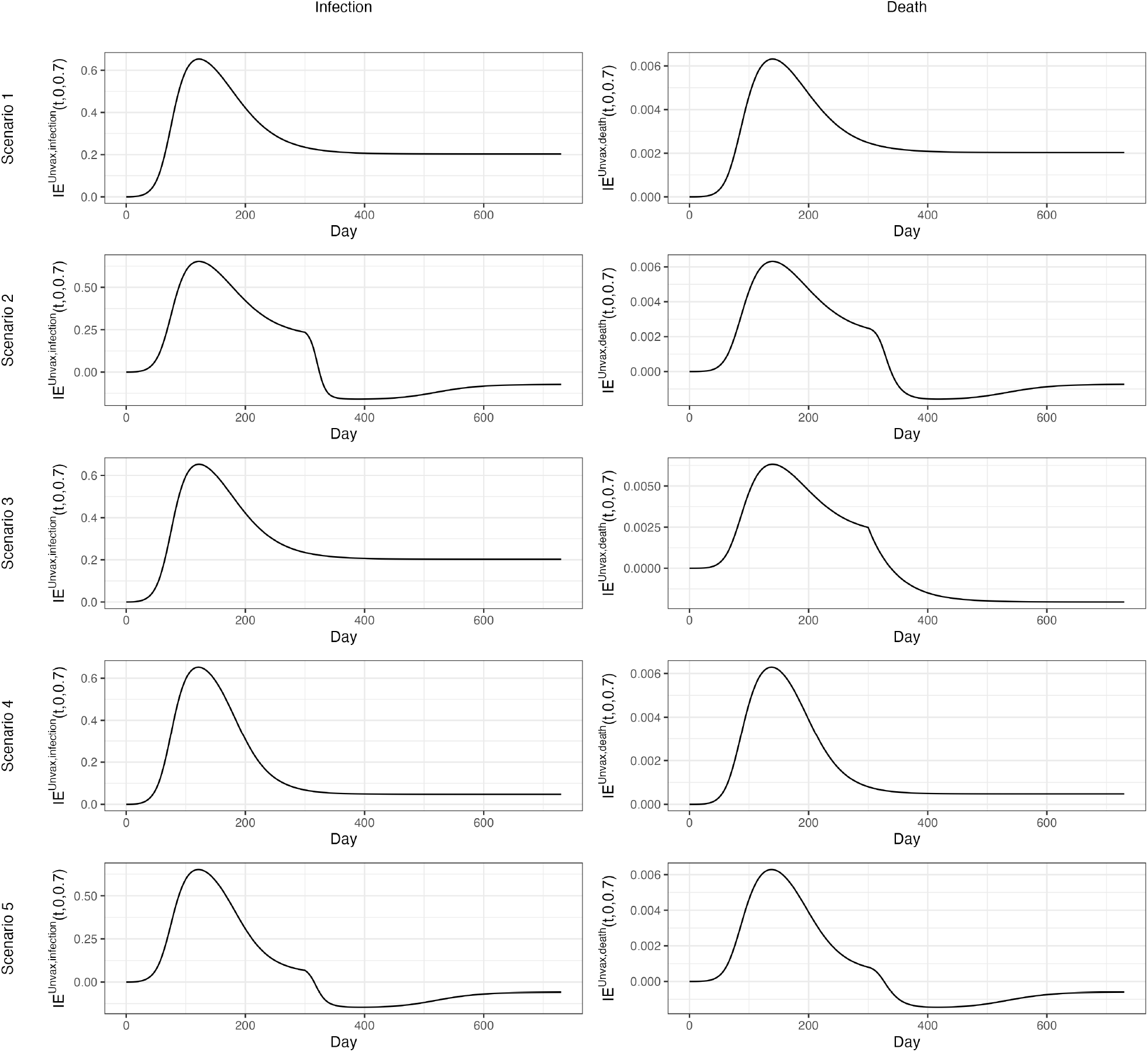
Indirect effect for the unvaccinated *IE*^*unvax*^(*t*, 0, 0. 7tblllll. Scenario 1, all parameters are time-invariant; Scenario 2, the number of effective contacts made by a typical infectious individual per day (*β*) increases from 0.15 to 0.6 at Day 300; Scenario 3, probability of infection-related death (*μ*) increases from 0.01 to 0.1 at Day 300; Scenario 4, vaccine efficacies against infection and death start to wane linearly after Day 100 reaching 0% at Day 300; and Scenario 5, the combination of Scenarios 2 and 4.

**eFigure 4.**
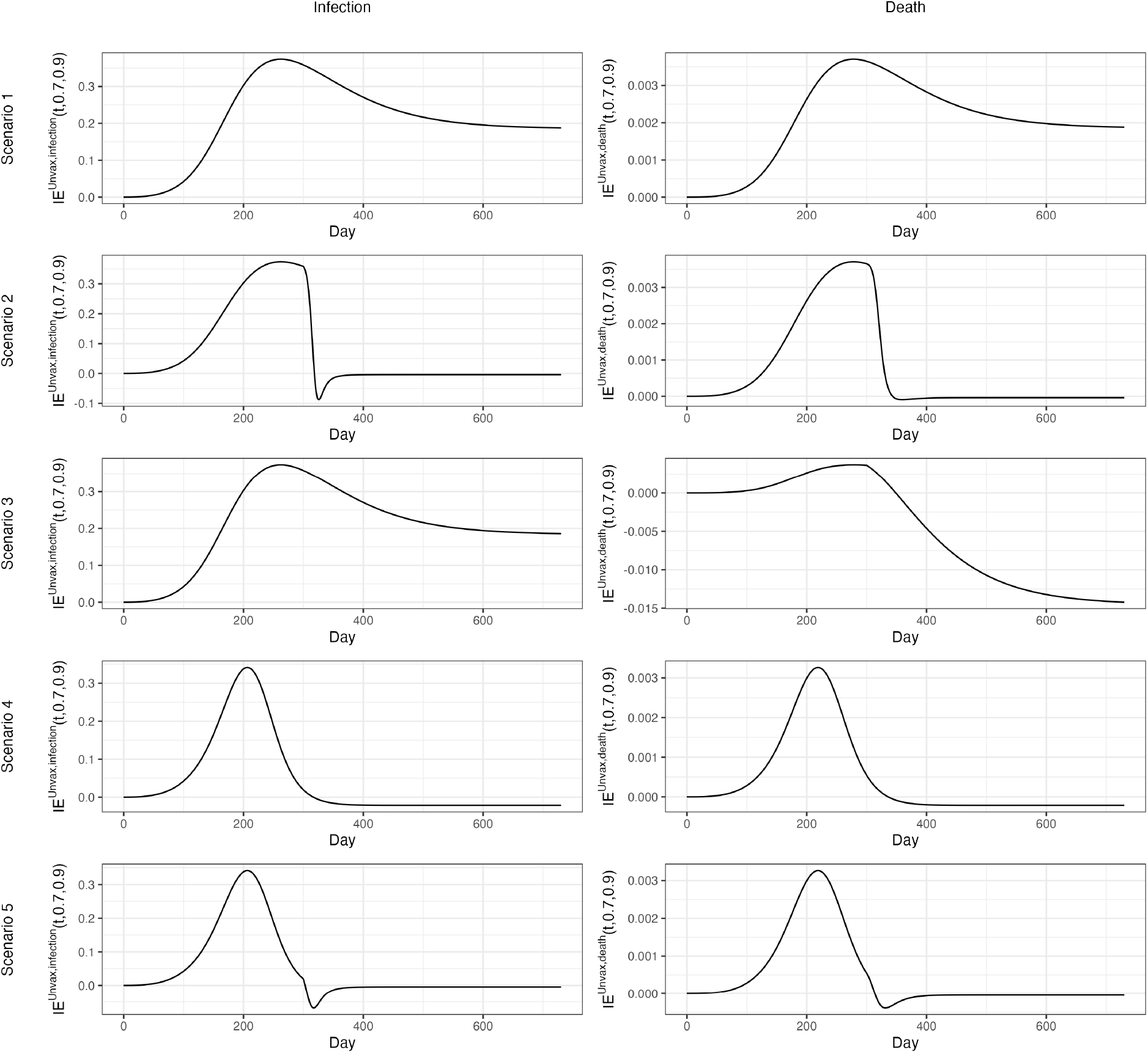
Indirect effect for the unvaccinated *IE*^*unvax*^(*t*, 0. 7, 0. 9).

**eFigure 5.**
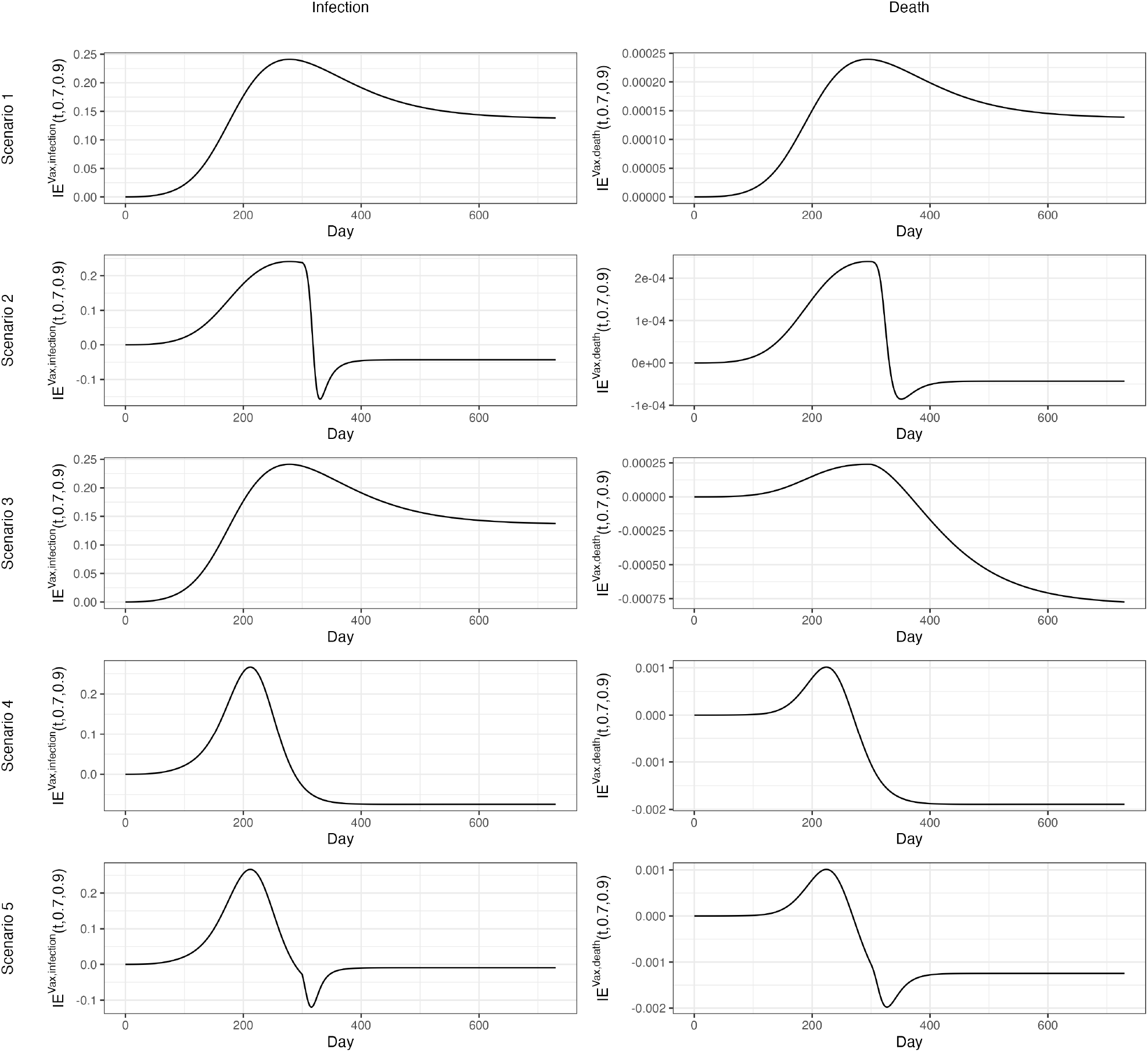
Indirect effect for the vaccinated *IE*^*vax*^(*t*, 0. 7, 0. 9).

### eAppendix 7. Trajectories of proportion susceptible by vaccination status over time

**eFigure 6.**
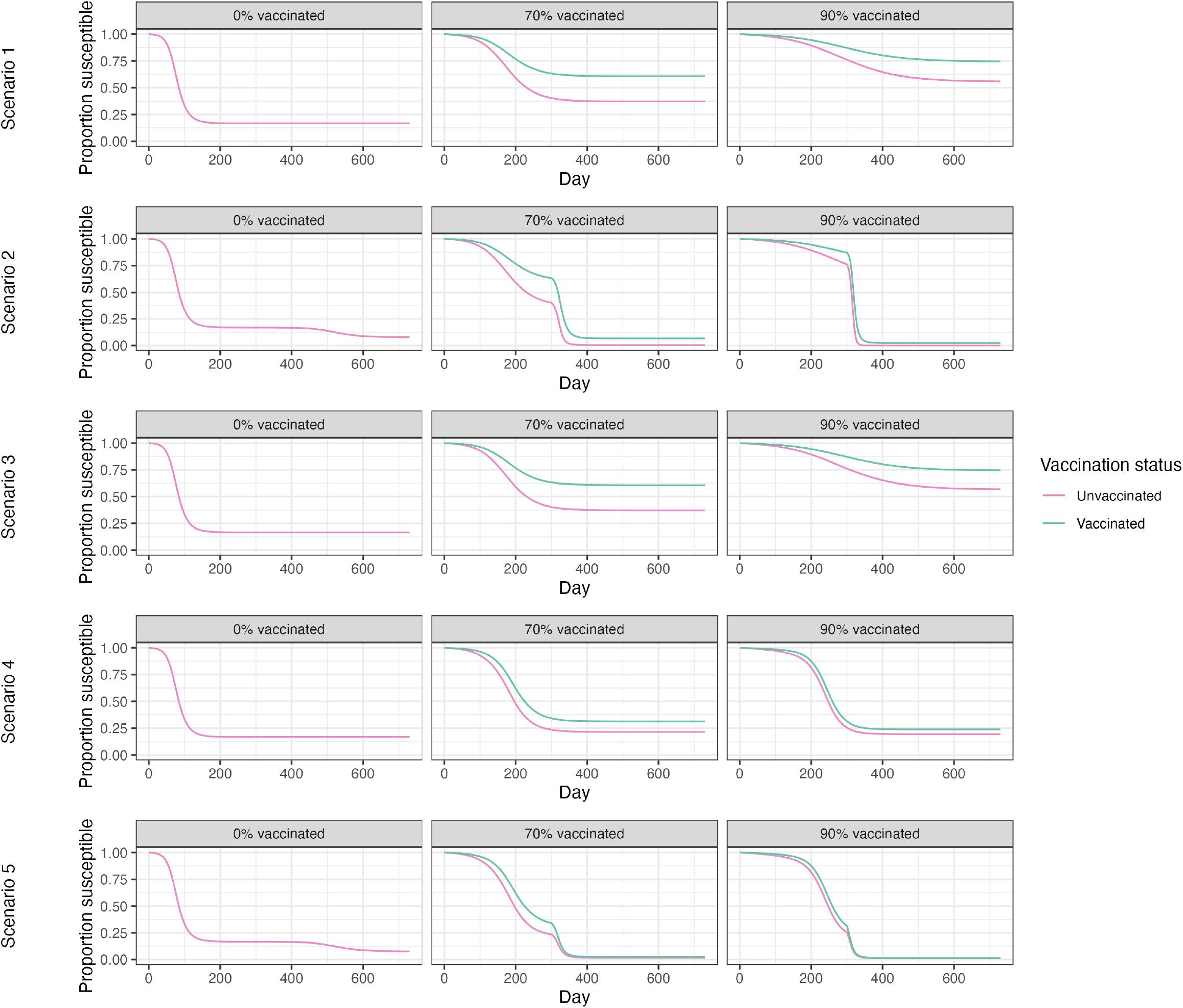
Proportion susceptible among vaccinated and unvaccinated individuals under 0%, 70%, or 90% proportions vaccinated. Scenario 1, all parameters are time-invariant; Scenario 2, the number of effective contacts made by a typical infectious individual per day (*β*) increases from 0.15 to 0.6 at Day 300; Scenario 3, probability of infection-related death (*μ*) increases from 0.01 to 0.1 at Day 300; Scenario 4, vaccine efficacies against infection and death start to wane linearly after Day 100 reaching 0% at Day 300; and Scenario 5, the combination of Scenarios 2 and 4.

### eAppendix 8. Epidemic curves under Scenario 4

eFigure 7 shows the epidemic curves given *α*_1_ = 0.7 and *α*_2_ = 0.9 proportions vaccinated. When *α*_1_ = 0.7, the epidemic peaks earlier and slows down due to the build-up of recovered individuals with sterilizing immunity. Consequently, return of full susceptibility (i.e., loss of protection) among vaccinated individuals is too late to rescue the epidemic. However, when *α*_2_ = 0.9, epidemic is delayed and a pool of individuals who are at risk of infection is built up, such that return of susceptibility among vaccinated individuals can rescue the epidemic.

**eFigure 7.**
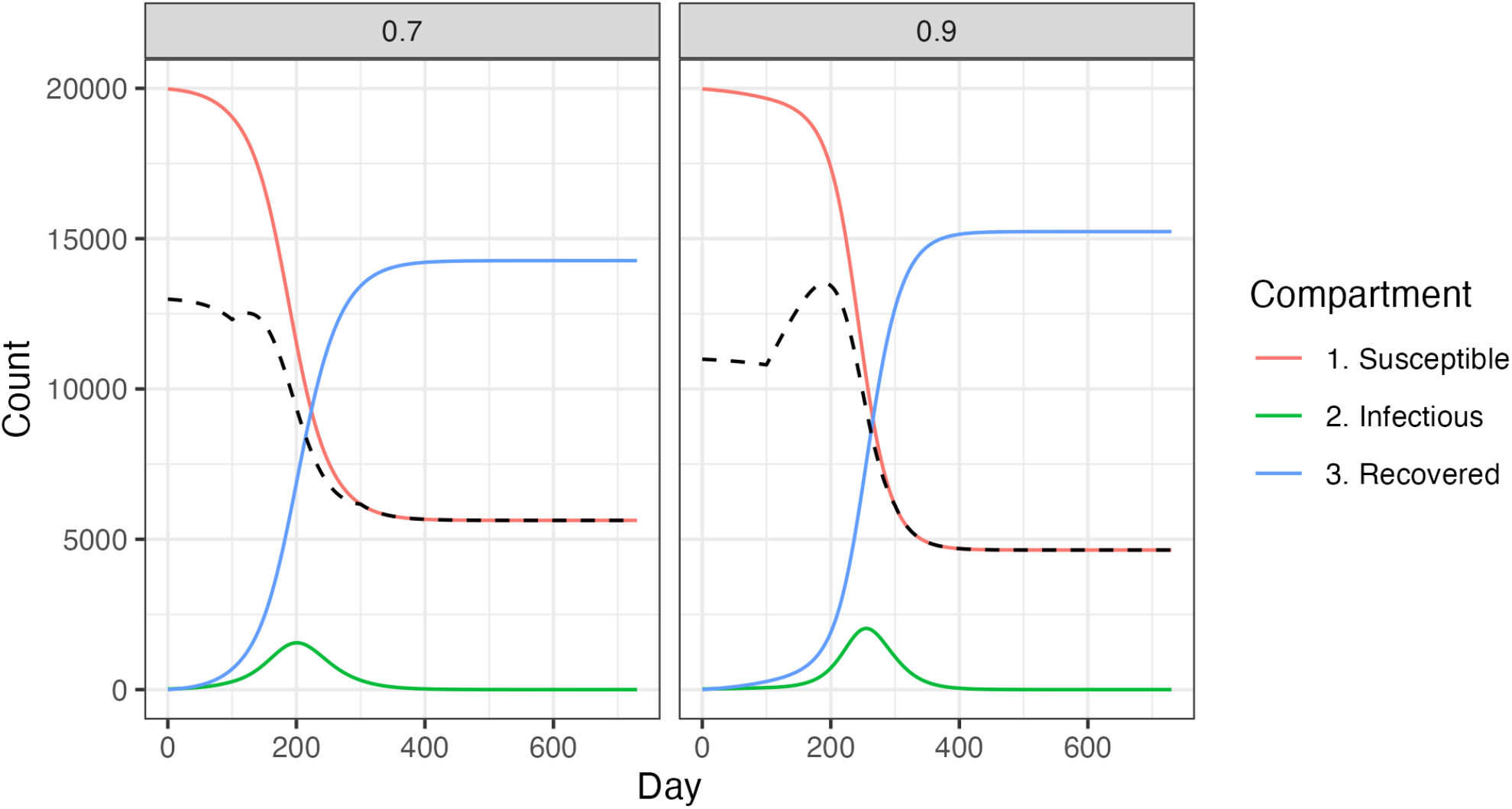
Epidemic curves under Scenario 4 given **α**_1_ = 0. 7 and **α**_2_ = 0. 9 proportions vaccinated. The dashed line represents the number of effectively susceptible persons = *S*_u_(*t*) + *θ*(*t*) ⋅ *S*_v_(*t*) where *S*_u_(*t*) is the number of unvaccinated susceptibles and *θ*(*t*) ⋅ *S*_v_(*t*) is the number of vaccinated susceptibles multiplied with *θ*(*t*) = 1 − *VE*_*infection*_ (*t*)/100%. The dashed lines show that more individuals at risk of infection (weighted by that risk) are built up under *α*_2_ = 0.9 than *α*_1_ = 0.7 before the epidemic peaks.

## REFERENCES

1. Vilches TN, Moghadas SM, Sah P, et al. Estimating COVID-19 Infections, Hospitalizations, and Deaths Following the US Vaccination Campaigns During the Pandemic. JAMA Netw Open. 2022;5(1):e2142725. doi:10.1001/jamanetworkopen.2021.42725

2. Schneider EC, Shah A, Sah P, et al. Impact of U.S. COVID-19 Vaccination Efforts: An Update on Averted Deaths, Hospitalizations, and Health Care Costs Through March 2022. To the Point. April 8, 2022. Accessed December 10, 2023. 10.26099/d3dm-fa91

3. Gavish N, Yaari R, Huppert A, Katriel G. Population-level implications of the Israeli booster campaign to curtail COVID-19 resurgence. Sci Transl Med. 2022;14(647):eabn9836. doi:10.1126/scitranslmed.abn9836

4. Haas EJ, McLaughlin JM, Khan F, et al. Infections, hospitalisations, and deaths averted via a nationwide vaccination campaign using the Pfizer–BioNTech BNT162b2 mRNA COVID-19 vaccine in Israel: a retrospective surveillance study. The Lancet Infectious Diseases. 2022;22(3):357–366. doi:10.1016/S1473-3099(21)00566-1

5. Brault A, Hart A, Uribe P, et al. Direct impact of COVID-19 vaccination in Chile: averted cases, hospitalizations, ICU admissions, and deaths. BMC Infect Dis. 2024;24(1):467. doi:10.1186/s12879-024-09304-1

6. Santos CVBD, Noronha TGD, Werneck GL, Struchiner CJ, Villela DAM. Estimated COVID-19 severe cases and deaths averted in the first year of the vaccination campaign in Brazil: a retrospective observational study. The Lancet Regional Health - Americas. 2023;17:100418. doi:10.1016/j.lana.2022.100418

7. Kayano T, Sasanami M, Kobayashi T, et al. Number of averted COVID-19 cases and deaths attributable to reduced risk in vaccinated individuals in Japan. The Lancet Regional Health - Western Pacific. 2022;28:100571. doi:10.1016/j.lanwpc.2022.100571

8. Jia KM, Hanage WP, Lipsitch M, et al. Estimated preventable COVID-19-associated deaths due to non-vaccination in the United States. Eur J Epidemiol. Published online April 24, 2023. doi:10.1007/s10654-023-01006-3

9. Zhong M, Glazer T, Kshirsagar M, et al. Estimating Vaccine-Preventable COVID-19 Deaths Among Adults Under Counterfactual Vaccination Scenarios in The United States: A Modeling Study Using Observational Data. J Pharm Pharmacol Res. 2023;07(03). doi:10.26502/fjppr.079

10. Halloran ME, Struchiner CJ. Study Designs for Dependent Happenings: Epidemiology. 1991;2(5):331–338. doi:10.1097/00001648-199109000-00004

11. VanderWeele TJ, Tchetgen Tchetgen EJ. Effect partitioning under interference in two-stage randomized vaccine trials. Statistics & Probability Letters. 2011;81(7):861–869. doi:10.1016/j.spl.2011.02.019

12. Hudgens MG, Halloran ME. Toward Causal Inference With Interference. Journal of the American Statistical Association. 2008;103(482):832–842. doi:10.1198/016214508000000292

13. Halloran ME, Struchiner CJ, Longini IM. Study Designs for Evaluating Different Efficacy and Effectiveness Aspects of Vaccines. American Journal of Epidemiology. 1997;146(10):789–803. doi:10.1093/oxfordjournals.aje.a009196

14. Polack FP, Thomas SJ, Kitchin N, et al. Safety and Efficacy of the BNT162b2 mRNA Covid-19 Vaccine. N Engl J Med. 2020;383(27):2603–2615. doi:10.1056/NEJMoa2034577

15. Frenck RW, Klein NP, Kitchin N, et al. Safety, Immunogenicity, and Efficacy of the BNT162b2 Covid-19 Vaccine in Adolescents. N Engl J Med. 2021;385(3):239–250. doi:10.1056/NEJMoa2107456

16. Standaert B, Dort T, Toumi M. Vaccine Efficacy, Effectiveness, Or Impact: Which One To Choose In Economic Evaluations Of Vaccines? Value in Health. 2017;20(9):A754. doi:10.1016/j.jval.2017.08.2115

17. Chakladar S, Rosin S, Hudgens MG, et al. Inverse probability weighted estimators of vaccine effects accommodating partial interference and censoring. Biometrics. 2022;78(2):777–788. doi:10.1111/biom.13459

18. Kayano T, Nishiura H. Assessing the COVID-19 vaccination program during the Omicron variant (B.1.1.529) epidemic in early 2022, Tokyo. BMC Infect Dis. 2023;23(1):748. doi:10.1186/s12879-023-08748-1

19. Watson OJ, Barnsley G, Toor J, Hogan AB, Winskill P, Ghani AC. Global impact of the first year of COVID-19 vaccination: a mathematical modelling study. The Lancet Infectious Diseases. 2022;22(9):1293–1302. doi:10.1016/S1473-3099(22)00320-6

20. Shoukat A, Vilches TN, Moghadas SM, et al. Lives saved and hospitalizations averted by COVID-19 vaccination in New York City: a modeling study. The Lancet Regional Health - Americas. 2022;5:100085. doi:10.1016/j.lana.2021.100085

21. Magosi LE, Zhang Y, Golubchik T, et al. Deep-sequence phylogenetics to quantify patterns of HIV transmission in the context of a universal testing and treatment trial – BCPP/Ya Tsie trial. eLife. 2022;11:e72657. doi:10.7554/eLife.72657

22. R Core Team. R: A Language And Environment for Statistical Computing. Accessed May 31, 2024. https://www.R-project.org/

23. FitzJohn R. odin: ODE Generation and Integration. Published online 2024. https://github.com/mrc-ide/odin

24. Shaman J, Kohn M. Absolute humidity modulates influenza survival, transmission, and seasonality. Proc Natl Acad Sci USA. 2009;106(9):3243–3248. doi:10.1073/pnas.0806852106

25. Li Y, Campbell H, Kulkarni D, et al. The temporal association of introducing and lifting non-pharmaceutical interventions with the time-varying reproduction number (R) of SARS-CoV-2: a modelling study across 131 countries. The Lancet Infectious Diseases. 2021;21(2):193–202. doi:10.1016/S1473-3099(20)30785-4

26. Simonsen L, Chowell G, Andreasen V, et al. A review of the 1918 herald pandemic wave: importance for contemporary pandemic response strategies. Annals of Epidemiology. 2018;28(5):281–288. doi:10.1016/j.annepidem.2018.02.013

27. Chemaitelly H, Tang P, Hasan MR, et al. Waning of BNT162b2 Vaccine Protection against SARS-CoV-2 Infection in Qatar. N Engl J Med. 2021;385(24). doi:10.1056/NEJMoa2114114

28. Menegale F, Manica M, Zardini A, et al. Evaluation of Waning of SARS-CoV-2 Vaccine–Induced Immunity: A Systematic Review and Meta-analysis. JAMA Netw Open. 2023;6(5):e2310650. doi:10.1001/jamanetworkopen.2023.10650

29. Lin L, Demirhan H, Stone L. Evaluating the Effectiveness of Vaccination Campaigns: A Sird-Type Model Versus a Statistical Approach. Published online 2024. doi:10.2139/ssrn.4940607

30. Pitman R, Fisman D, Zaric GS, et al. Dynamic Transmission Modeling: A Report of the ISPOR-SMDM Modeling Good Research Practices Task Force Working Group–5. Med Decis Making. 2012;32(5):712–721. doi:10.1177/0272989X12454578

31. Leung NHL, Chu DKW, Shiu EYC, et al. Respiratory virus shedding in exhaled breath and efficacy of face masks. Nat Med. 2020;26(5):676–680. doi:10.1038/s41591-020-0843-2

32. Bushman M, Kahn R, Taylor BP, Lipsitch M, Hanage WP. Population impact of SARS-CoV-2 variants with enhanced transmissibility and/or partial immune escape. Cell. 2021;184(26):6229–6242.e18. doi:10.1016/j.cell.2021.11.026

33. Horwitz LI, Jones SA, Cerfolio RJ, et al. Trends in COVID-19 Risk-Adjusted Mortality Rates. Journal of Hospital Medicine. 2021;16(2):90–92. doi:10.12788/jhm.3552

34. Dennis JM, McGovern AP, Vollmer SJ, Mateen BA. Improving Survival of Critical Care Patients With Coronavirus Disease 2019 in England: A National Cohort Study, March to June 2020. Critical Care Medicine. 2021;49(2):209–214. doi:10.1097/CCM.0000000000004747

35. Panagiotopoulos T, Antoniadou I, Valassi-Adam E. Increase in congenital rubella occurrence after immunisation in Greece: retrospective survey and systematic review. 1999;319.

36. Markov PV, Ghafari M, Beer M, et al. The evolution of SARS-CoV-2. Nat Rev Microbiol. 2023;21(6):361–379. doi:10.1038/s41579-023-00878-2

## References for eAppendix

1. VanderWeele TJ, Tchetgen Tchetgen EJ. Effect partitioning under interference in two-stage randomized vaccine trials. Statistics & Probability Letters. 2011;81(7):861–869. doi:10.1016/j.spl.2011.02.019

2. Hudgens MG, Halloran ME. Toward Causal Inference With Interference. Journal of the American Statistical Association. 2008;103(482):832–842. doi:10.1198/016214508000000292

3. FitzJohn R. odin: ODE Generation and Integration. Published online 2024. https://github.com/mrc-ide/odin

4. Soetaert K, Petzoldt T R. Woodrow S. Solving Differential Equations in R: Package deSolve. Journal of Statistical Software. 2010;33(9):1–25. doi:10.18637/jss.v033.i09

5. Miller JC. A Note on the Derivation of Epidemic Final Sizes. Bull Math Biol. 2012;74(9):2125–2141. doi:10.1007/s11538-012-9749-6

6. Lin L, Hamedmoghadam H, Shorten R, Stone L. Quantifying indirect and direct vaccination effects arising in the SIR model. J R Soc Interface. 2024;21(218):20240299. doi:10.1098/rsif.2024.0299

7. Chao DL, Dimitrov DT. Seasonality and the effectiveness of mass vaccination. Mathematical Biosciences and Engineering. 2016;13(2):249–259. doi:10.3934/mbe.2015001

8. Carnell R. lhs: Latin Hypercube Samples. Published online 2022. https://CRAN.R-project.org/package=lhs

